# Unmasking the J-wave: 3D ECG shows terminal depolarization mimicking early repolarization

**DOI:** 10.64898/2025.11.30.25341283

**Authors:** Alejandro Jesús Bermejo Valdés

**Affiliations:** Riojan Health Service, Logroño, Spain

**Keywords:** Three-dimensional ECG, J-wave, Early repolarization, 3D loops

## Abstract

We used three-dimensional (3D) electrocardiography (ECG) to track the J-wave in V6 through viewing planes inaccessible to standard ECG. Across three identified J-loop phenotypes, we found that the planar morphology of the end QRS deflection, whether J-wave or slurring, depended on projection and was interconvertible under rotation. Although traditionally attributed to phase-1 of the action potential, the 3D J-loop showed that the J-wave becomes concealed and occupies depolarization regions in V1. Here, we examine this discrepancy and show that 3D ECG can more accurately reinterpret the electrophysiological and pathological phenomena associated with early repolarization.

## 1. Introduction

The J-wave, classically defined as a positive deflection at the end of the QRS complex, is observed in ion channelopathies such as early repolarization (ER) and Brugada syndrome (BrS), as well as in metabolic or environmental conditions including hypercalcemia or hypothermia, and is associated with arrhythmogenic vulnerability and sudden cardiac death [1]. Experimental work has attributed its cellular and molecular basis to the I_to_ channels of cardiomyocytes from different myocardial layers, whose differential expression generates the phase-1 notch of the ventricular action potential [2, 3]. Despite this mechanistic insight, the diagnostic interpretation of the J-wave remains challenging because the terminal QRS complex and the onset of the ST segment constitute a transitional zone in which late depolarization and the initial phase of repolarization may coexist, a circumstance that also accounts for end QRS slurring as an additional manifestation of ER [1], reflecting a subtle overlap between these two phases of the cardiac cycle. The ambiguity inherent to this boundary region, where equivalent electrocardiographic (ECG) morphologies (J-wave and end QRS slurring) can arise in the context of ER, underscores the need for a more precise physiological interpretation.

In principle, the presence of a J-wave should be traceable across leads, maintaining a coherent spatial position. It is physiologically inconsistent to observe a J-wave at the onset of the ST segment in one lead while its corresponding points appear embedded within the QRS complex in another lead. However, the conventional 12-lead ECG provides only a restricted set of two-dimensional (2D) projections of cardiac electrical activity, thereby precluding continuous visualization of the spatial relationships between leads.

Our recent work described how standard leads can be combined to reconstruct a continuous three-dimensional (3D) electrical trajectory that preserves spatial relationships between leads [4]. This approach allows deflections to be tracked across intermediate viewing planes [5], thereby revealing projection-dependent changes that remain inaccessible to conventional 2D ECG analysis. Here, using this 3D method, we evaluate how terminal QRS deflections migrate and transform across precordial orientations and identify distinct phenotypes of J-wave behavior.

## 2. Methods

Signals were obtained from the PTB Diagnostic ECG Database [6]. We selected patients with a J-wave or QRS slurring in V6. For very small J-waves or slurring, we relied on the contiguous lead V5 and required a visible J-wave. Recordings with excessive noise or strictly filiform 3D loops (tightly clustered limbs), which prevent reliable geometric tracking [5], were excluded. The final dataset consisted of 31 patients.

For each selected recording, we extracted the first complete cardiac cycle occurring within the initial 2000 ms of the trace, sampled at 1000 Hz. Signal preprocessing was performed in Python 3.13.3 using the `NumPy`, `WFDB`, and `Matplotlib` libraries. A 0.5 Hz high-pass Butterworth filter was applied to remove baseline drift, and a 40 Hz low-pass Butterworth filter was used to suppress high-frequency noise while preserving J-wave morphology. Signal quality and spectral content were verified to be consistent with conventional clinical filtering (50 Hz notch; 0.05-150 Hz bandwidth).

3D ECG reconstruction followed the approach outlined in our recent work [4, 5]. We employed the *Trivial Representation*, embedding the precordial signals into a 3D space through the time-parameterized vector **p**(*t*) = (*V* 1(*t*), *V* 6(*t*), *t*). This yields a continuous trajectory *γ*(*t*) *⊂* ℝ^3^ that incorporates intermediate electrical information between leads V1 and V6, whose projections correspond to planar sections of a higher-dimensional ECG model. The resulting trajectories were rendered using mplot3d, and the J-wave segment was manually annotated in red.

All high-definition PNG figures, code, patient-specific time windows, the patient-by-patient summary of wave morphologies (J-wave or slurring) with their rotational transformations, and the list of excluded cases are available in the Mendeley Data repository [7]. The supplementary material accompanying this article provides a clinically focused subset, consisting of two PDF documents: one with all sample-specific 3D ECG figures (supplementary_3DECG_figures.pdf) and another summarizing the wave morphologies (supplementary_classification.pdf).

## 3. Results

Three J-loop phenotypes emerged from 3D tracking of the terminal QRS deflection across precordial orientations, which we classified into three groups (groups *i*-*iii*). In addition, we identified bidirectional slurring-to-J-wave interconversion under small rotational adjustments around the time axis.

### 3.1. Group i: Concealed J-loop unmasked by 3D rotation

In these recordings, the 3D J-loop corresponding to the 2D J-wave or slurring in V6 was not visible in the V1 projection and became apparent only after rotation of the 3D ECG (Fig. 1). A total of 25 cases (80.6 %) presented this configuration.

**Figure 1:**
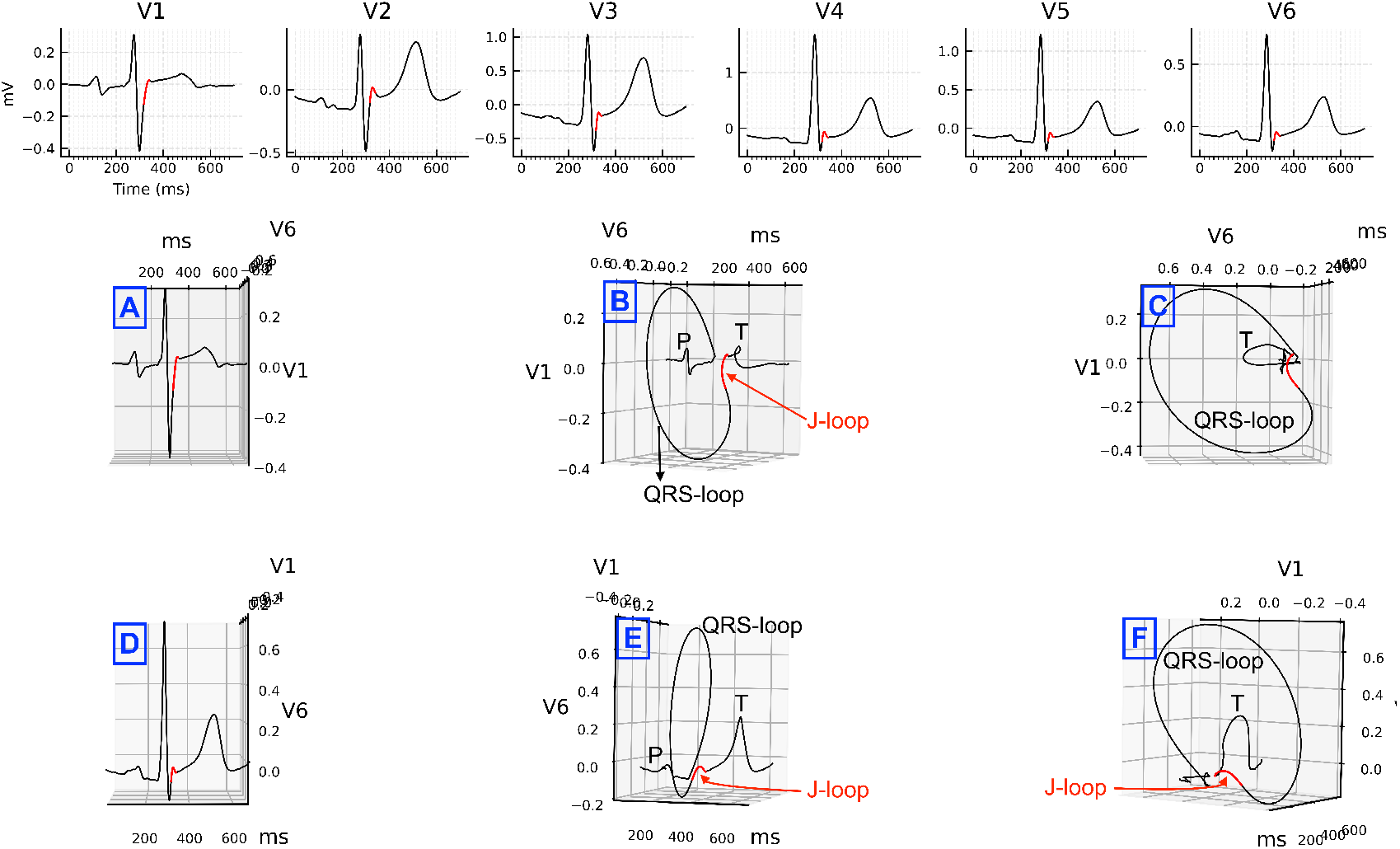
Group *i*: Representative example (patient 180). Top row: precordial leads V1-V6. Middle row: 3D ECG loops obtained from sequential leftward rotations starting from the V1-oriented view (A, B and C); the rightmost panel (C) shows the fully lateral V1-V6 projection. Bottom row: projections centered on the V6 orientation (D, frontal; E, leftward-rotated; F, rightward-rotated). Selected projections include labels marking major loops and waves (QRS-loop, J-loop, P-wave, and T-wave). The J-loop is highlighted in red. The J-wave is concealed in the V1 projection and becomes progressively unmasked as the 3D loop is rotated.

The cases included (patient/record) were: 017/s0053lre, 040/s0219lre, 045/s0147lre, 048/s0171lre, 049/s0173lre, 052/s0190lre, 057/s0198lre, 083/s0268lre, 099/s0387lre, 110/s0003_re, 165/s0322lre, 173/s0305lre, 180/s0374lre, 198/s0402lre, 240/s0468_re, 241/s0469_re, 245/s0474_re, 248/s0481_re, 249/s0484_re, 250/s0485_re, 252/s0487_re, 258/s0494_re, 259/s0495_re, 273/s0511_re, and 277/s0527_re.

### 3.2. Group ii: J-loop polarity symmetry

In this phenotype, the J-wave in V6 remains spatially linked to a small terminal QRS deflection in V1 with the same polarity, irrespective of the V1-V6 3D rotation (Fig. 2). We refer to this behavior as a polarity symmetry of the J-loop under global rotation (3D system and its embedded function) around the time axis. This group includes the following cases (9.7 %): 043/s0141lre, 230/s0454_re, and 244/s0473_re.

**Figure 2:**
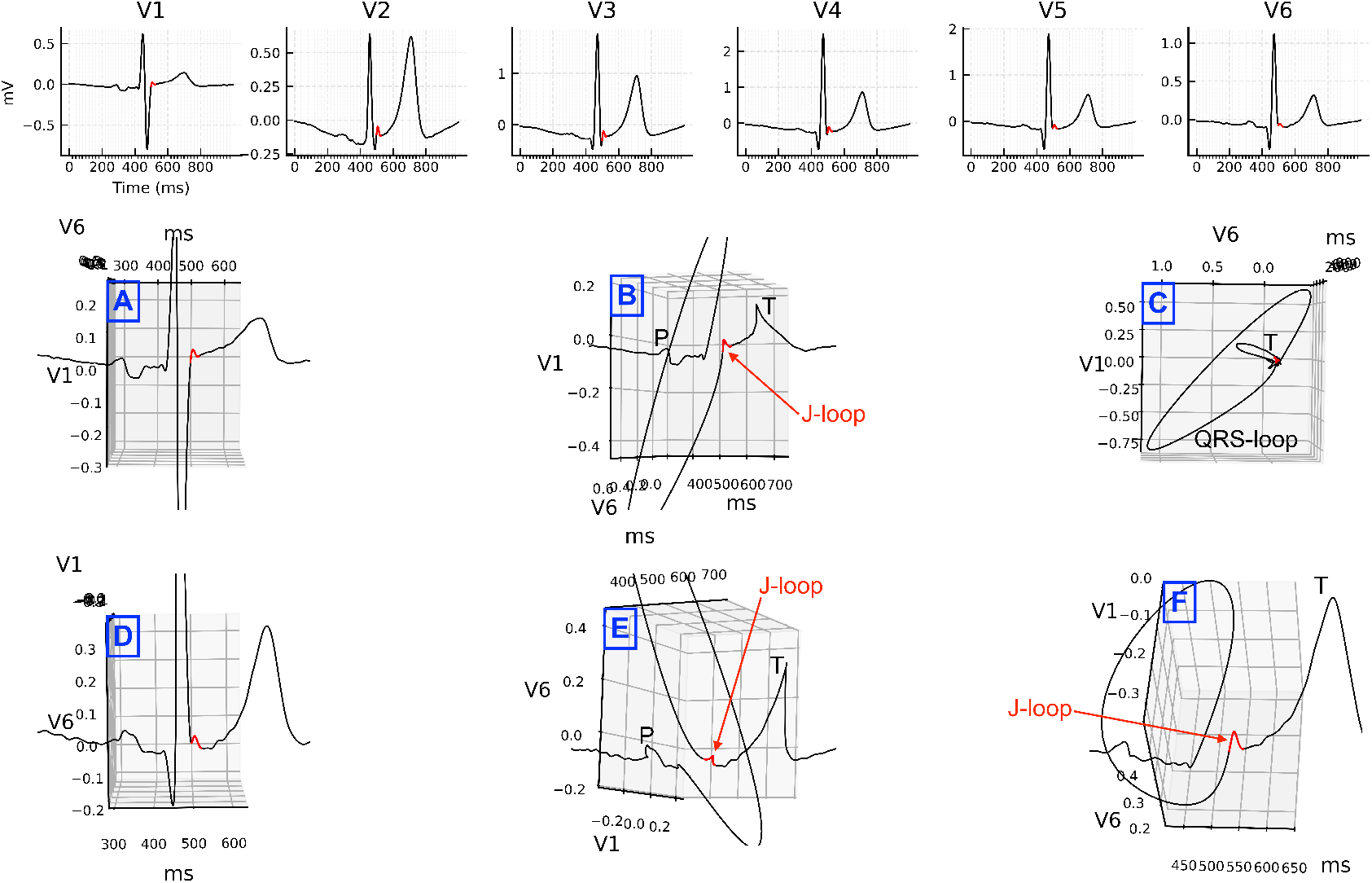
Group *ii*: Representative example (patient 244). Top row: precordial leads V1-V6. Middle row: 3D projections obtained from the V1-oriented view (A), a leftward rotation (B), and the fully lateral V1-V6 plane (C). Bottom row: additional projections derived from the V6 orientation (D), including upper-left (E) and lower-left (F) rotated views. Selected panels include annotations marking major loop structures (QRS-loop and J-loop) as well as the P- and T-wave. A slight zoom was applied to the 3D views to improve visualization. The J-loop (highlighted in red) preserves its positive polarity from V1 through the V6-oriented projections.

### 3.3. Group iii: J-loop polarity inversion

In this group (Fig. 3), the configuration resembles that of group *ii* but with polarity inversion: a small negative terminal QRS deflection in V1 becomes a J-wave as the 3D rotation approaches the V6 orientation. The recordings that exhibited this behavior are (9.7 %): 073/s0238lre, 150/s0287lre, and 187/s0207_re.

**Figure 3:**
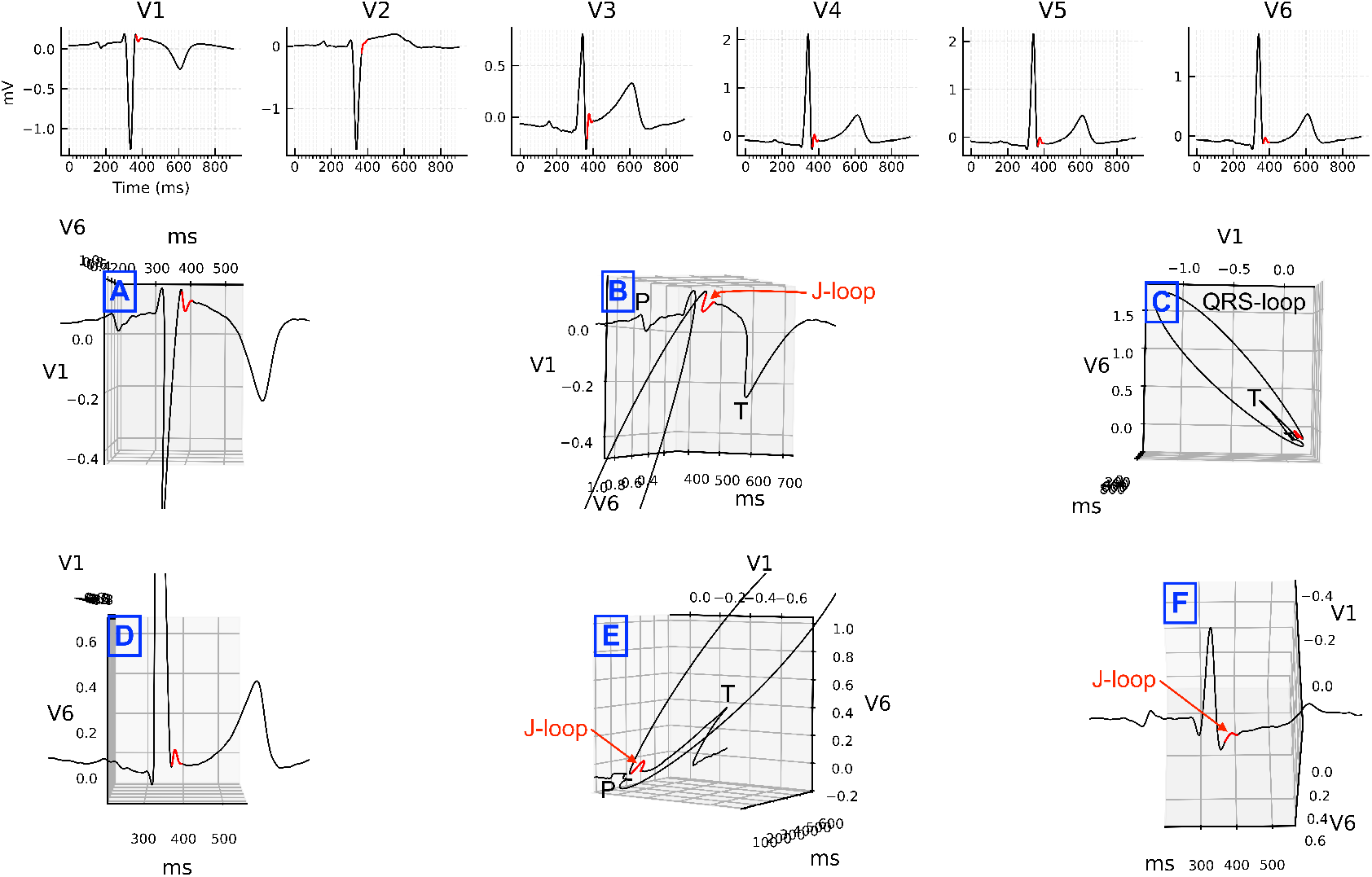
Group *iii*: Representative example (patient 150). Top row: precordial leads V1-V6. Middle row: V1-oriented view (A), a leftward rotation from the V1 orientation (B), and the fully lateral V1-V6 plane (C). Bottom row: V6-oriented view (D), a rightward rotation (E), and a hybrid intermediate view between V1 and V6 (F). Selected panels include annotations marking major loop structures (QRS-loop and J-loop), as well as the P- and T-wave. A slight zoom was applied to the 3D views to improve visualization. The J-loop (highlighted in red) exhibits polarity inversion along the V1-to-V6 trajectory.

### 3.4. Slurring-J-wave interconversion under 3D rotation

Rotation of the 3D ECG system from a V6-oriented view toward hybrid V1-V6 projections revealed group-specific interconversion patterns. Among patients with a J-wave in V6 (20 in group *i*, 3 in group *ii*, and 2 in group *iii*), J-wave *→* slurring transitions were observed exclusively in group *i* under 3D rotation. No such transitions occurred in groups *ii* or *iii*. In contrast, all patients exhibiting slurring in V6 (5 in group *i*: 045, 057, 083, 110, and 273; and 1 in group *iii*: 187) developed a J-wave after rotational adjustments of the 3D viewing space (example in Appendix A.1) (see supplementary material/dataset [7] for all figures and the patient-by-patient morphology classification and rotational transformations).

## 4. Discussion

Across the three phenotypes examined, we observed that the same J-loop can exhibit different morphologies depending on the viewing plane. In the first and most frequent group (*≈*81 %), the J-wave observed in V6 is embedded within the terminal QRS in V1. Only the 3D reconstruction reveals its continuity, which appears as a J-loop concealed behind the V1 projection. In the second group, a symmetric J-loop preserves its polarity during the V1-V6 rotation, whereas in the third group a polarity inversion appears. These patterns indicate that the presentation of the J-wave morphology, whether in its shape, position, or inversion, depends on the J-loop geometry within the terminal QRS region.

Our observations, largely derived from group *i*, may refine the conventional understanding of the J-wave by showing that a deflection located at the J-position cannot be assumed to reflect the initial phases of repolarization unless its spatial coherence across leads is demonstrated. If the J-wave is interpreted as a manifestation of repolarization, then the terminal portion of the QRS in V1 must also be regarded as being influenced by repolarization. Conversely, if the terminal QRS in V1 represents a depolarization phenomenon, then its projection in V6 (manifesting as the J-wave) must likewise arise from depolarization. The planar ECG lacks the geometric information needed to resolve this ambiguity, particularly in defining J-loop continuity and morphology, making 3D tracking essential for characterizing its topology across the leads.

Our results support four complementary interpretations of the J-wave:

### 4.1. The J-wave as an ER phenomenon

If the J-wave indeed marks the onset of repolarization in V6, the terminal portion of the QRS in V1 must be regarded as reflecting the same repolarization process. This view is consistent with the molecular model in which the transient outward current I_to_ generates the phase-1 notch underlying ER patterns [1–3]. In this context, a J-loop concealed behind the QRS in V1 suggests that the terminal QRS in certain leads may reflect repolarization rather than late depolarization. This framework also aligns the slurred terminal QRS in V6 with the terminal QRS in V1, which is reasonable given that both represent intrinsic components of the QRS. However, the electrophysiological correspondence between the J-wave in V6 and the terminal QRS in V1 is more difficult to explain.

### 4.2. The J-wave as late depolarization

Our data also support an alternative mechanism, opposite to the former, in which the J-wave arises from terminal depolarization rather than from the initial phase of repolarization. This model is compatible with clinical observations describing J-wave appearance in diseases with conduction delay such as non-compaction cardiomyopathy, arrhythmogenic right ventricular cardiomyopathy, and myocardial ischemia [8].

The proposal that the J-wave may arise from intraventricular conduction delays is known as the depolarization hypothesis, which assumes that these delays occur in the terminal portion of the QRS complex, thereby masquerading as a repolarization-related deflection [1]. Although these mechanisms have been developed largely in the context of BrS, ER shares a related electrophysiological substrate, which has facilitated the transfer of conceptual models between both entities. Their foundations, however, are not identical. Anatomically, BrS primarily involves the anterior right ventricular outflow tract (RVOT), whereas ER is most often linked to the inferior left ventricular region [1], indicating distinct spatial substrates for J-wave formation. In fact, the depolarization-based framework is rooted in RVOT pathology [3], which limits its applicability to ER, and there is molecular evidence supporting this view. For example, sodium channel blockers unmask or accentuate J-wave manifestations in BrS but reduce their amplitude in ER [2], further underscoring their divergent underlying mechanisms despite surface ECG similarity.

### 4.3. The J-wave as a depolarization-repolarization overlap zone

A third possibility is that the J-wave reflects a transitional zone in which late depolarization coexists with phase-1, as a result of temporal dispersion in depolarization and repolarization across different myocardial regions. Anatomical spacing and cellular heterogeneity across myocardial layers may allow repolarization to begin in one region while depolarization continues in another, both contributing to the same surface deflection. The coexistence of these processes within the terminal QRS may explain why slurring in ER often merges seamlessly with the end of depolarization. In this framework, the J-loop may represent the geometric locus where the terminal portion of depolarization and the onset of repolarization converge in 3D space. It is plausible that phase-0 and phase-1 operate in an interdependent manner at the molecular level, producing a unified signature on the surface ECG that becomes apparent within the J-loop. In this view, the J-loop reflects a coupled depolarization-repolarization interface rather than the isolated expression of a single component. Experimental evidence supports this framework, showing that the ionic processes governing these phases can modulate one another through shared regulatory and intracellular signaling mechanisms [3], leading to a hybrid manifestation on the 3D ECG.

### 4.4. The J-wave and slurring as planar manifestations of the J-loop

The relationship between end QRS slurring and the J-wave in contiguous leads remains uncertain. In our cohort, all six patients who presented slurring in V6 displayed a J-wave after rotation of the 3D ECG system around the time axis. Conversely, patients in group *i* with a J-wave in V6 also showed J-wave *→* slurring interconversion under rotation; however, this direction of morphological change appears trivial compared with the slurring *→* J-wave transition. These findings suggest that slurring and the J-wave may represent different planar manifestations of the same underlying 3D trajectory.

A similar phenomenon has been reported in standard 2D ECG by Macfarlane and co-workers [9], who described a J-wave in V4 evolving into end QRS slurring in V6. Within our framework, such evolution is readily explained by progressive rotations through the 3D ECG space, highlighting that terminal QRS morphology is inherently projection dependent (Appendix A.1).

Stratification by phenotypic groups further refined this interpretation. In group *i*, all patients exhibited fully bidirectional interconversion. By contrast, groups *ii* and *iii* were largely deficient in this regard, with J-waves remaining morphologically stable under rotation and only the case presenting slurring in V6 displaying a J-wave configuration after rotation. These observations suggest that the ability to transition between J-wave and slurring is not uniform across phenotypes and may reflect subtle differences in the underlying 3D loop geometry and its projection.

The presence of a J-wave or end QRS slurring is a necessary, although not sufficient, criterion for the ER pattern, which additionally requires that either the peak of the J-wave or the onset of the slurring reaches at least 0.1 mV in *≥*2 contiguous leads (excluding V1-V3), with a QRS *<*120 ms [1]. However, our analysis indicates that the amplitude, morphology, and even the apparent presence of the J-wave depend on the projection plane of a higher-dimensional ECG representation and can be tracked throughout the entire 3D ECG space. Consequently, the slurring onset in V6 may exceed the J-wave peak amplitude under a slightly rotated projection (Appendix A.1), even though both manifestations arise from the same underlying phenomenon.

## 5. Conclusions

Our findings show that the underlying mechanism, whether delayed depolarization or initial repolarization, does not alter the geometric observation: the J-loop unifies the terminal QRS, J-waves, and slurring as distinct planar projections of a single 3D structure. From this perspective, the J-loop emerges as a key integrative element, providing a point of convergence between depolarization- and repolarization-based explanations.

The surface J-wave should not be regarded as a fixed electrophysiological entity but rather as the planar expression of a 3D electrical trajectory whose appearance depends on geometric context. Within this framework, 3D ECG reconstruction offers not only a means to visualize J-loops and their morphology but also a conceptual tool for refining the geometric interpretation of the J-wave, an aspect that merits further experimental and clinical investigation.

The clinical implications of these findings are considerable. J-waves and end QRS slurring are established markers of arrhythmic vulnerability, traditionally interpreted on the basis of their morphology and quantitative features. However, our identification of the underlying J-loop shows that these parameters vary as a function of the projection plane within the 3D ECG model. If so, current prognostic interpretations may require reevaluation. The 3D architecture of the J-loop could provide a more coherent substrate for risk stratification in ER.

## Supporting information

Patient-level 3D ECG figures grouped by J-loop phenotype, showing planar leads and rotated 3D trajectories for all included cases.

Patient-by-patient classification of V6 terminal QRS deflections and the corresponding rotational transformations, including excluded cases.

## Data Availability

All ECG recordings used in this study are publicly available in the PTB Diagnostic ECG Database at PhysioNet. All derived data products, 3D ECG reconstructions, and analysis scripts are available in the Mendeley Data repository referenced in the manuscript.

## Appendix A. Slurring-to-J-wave transition

### Appendix A.1. Recording 045

**Figure A.4:**
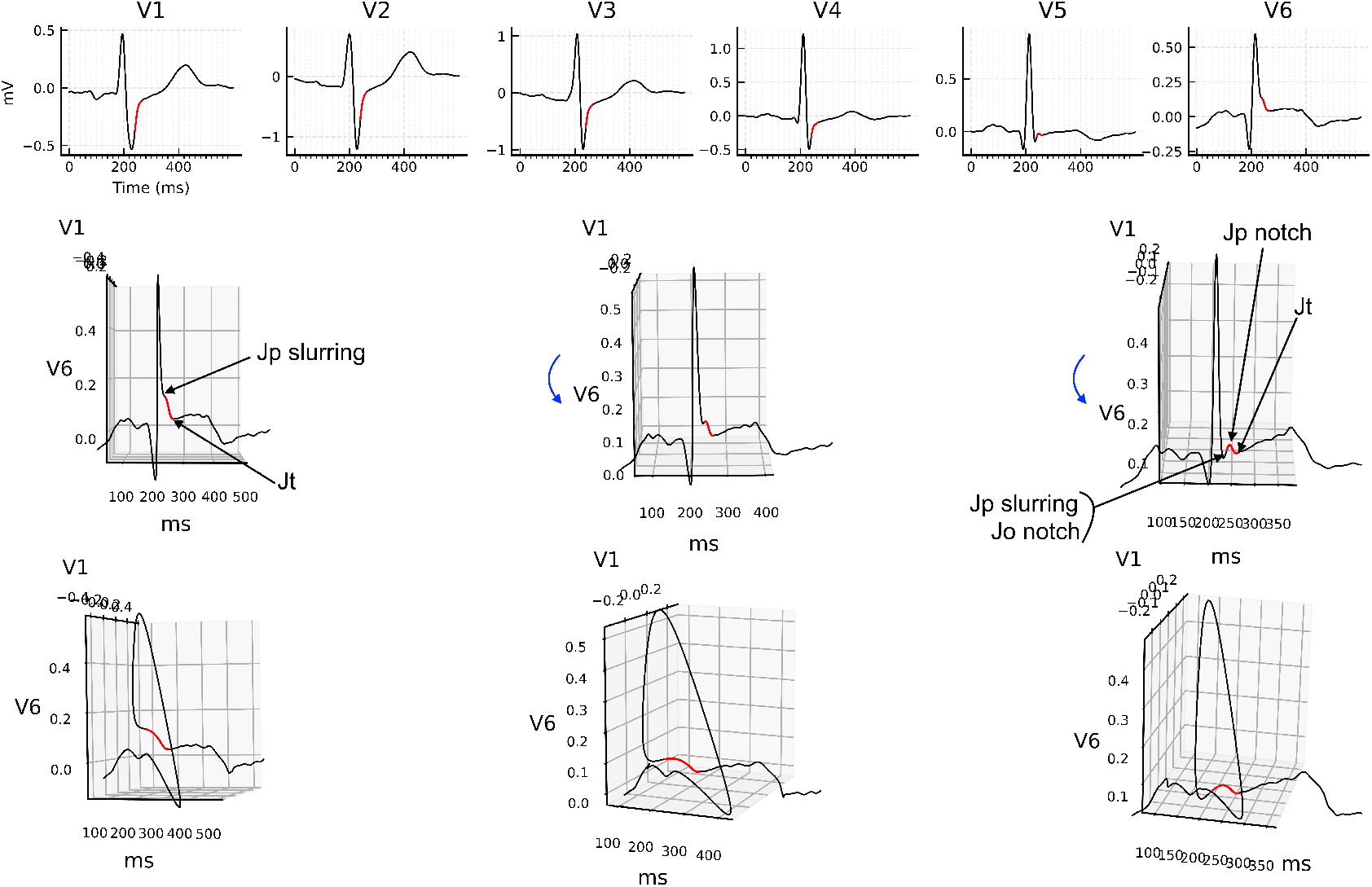
3D reconstruction of recording 045 showing the slurring-to-J-wave transition. Top row: precordial leads V1-V6 with the J-region highlighted in red. Middle row: 3D ECG loop viewed from a V6-oriented projection, where the onset of slurring (Jp slurring), the onset of the J-wave (Jo notch), the peak of the J-wave (Jp notch), and the termination of both the J-wave and the slurring (Jt) are annotated. Curved blue arrows indicate the direction of the applied 3D rotation. Bottom row: left-rotated projections of the middle row view. Note that Jp slurring and Jo notch coincide under slight anterior rotation, making their Jp/Jo-Jt intervals identical because Jt also aligns for both. In contrast, Jp notch does not coincide with Jp slurring in this projection. The notation is consistent with that used by Macfarlane and co-workers [9].

## Notes

### Competing Interest Statement

The authors have declared no competing interest.

### Funding Statement

This study did not receive any funding.

### Author Declarations

The PTB Diagnostic ECG Database, available at PhysioNet (https://physionet.org/content/ptbdb/1.0.0/). All recordings are fully anonymized and openly accessible without registration or data-use agreements.

### Summary of Updates

This version of the manuscript includes expanded methodological details, additional patient cases, and new descriptions of the three J-loop phenotypes and their rotational behavior. The supplementary material has been extended to include patient-level figures, morphology summaries, and the time windows used in the analysis, facilitating reproducibility. Minor editorial revisions were made throughout the text to enhance consistency with the updated content.

## References

[1] J-wave syndromes expert consensus conference report: Emerging concepts and gaps in knowledge, Heart Rhythm 13 (10) (2016) e295–e324, focus Issue: Sudden Death. doi:10.1016/j.hrthm.2016.05.024.

[2] C. Antzelevitch, G.-X. Yan, J-wave syndromes: Brugada and early repolarization syndromes, Heart Rhythm 12 (8) (2015) 1852–1866. doi: 10.1016/j.hrthm.2015.04.014.

[3] C. Antzelevitch, J. M. Di Diego, J wave syndromes: What’s new?, TRENDS. CARDIOVAS. MED. 32 (6) (2022) 350–363. doi:10.1016/j.tcm.2021.07.001.

[4] A. J. Bermejo Valdés, Three-dimensional standard electrocardiogram: A first approach based on precordial leads, J. Electrocardiol. 89 (2025) 153875, epub 2025 Jan 20. PMID: 39892036. doi:10.1016/j.jelectrocard.2025.153875.

[5] A. J. Bermejo Valdés, R-wave progression or r-wave rotation? a 3d ecg-based perspective, J. Electrocardiol. 91 (2025) 154047, epub 2025 Jun 7. PMID: 40513181. doi:10.1016/j.jelectrocard.2025.154047.

[6] R. Bousseljot, D. Kreiseler, A. Schnabel, Nutzung der ekg-signaldatenbank cardiodat der ptb über das internet, Biomed. Eng. 40 (Suppl 1) (1995) 317. URL https://physionet.org/content/ptbdb/1.0.0/

[7] A. J. Bermejo Valdés, 3d ecg: J-wave groups dataset (2025). doi: 10.17632/myhw9rcjp3.2.

[8] A. S. Tsvetkova, J. E. Azarov, O. G. Bernikova, A. O. Ovechkin, M. A. Vaykshnorayte, M. M. Demidova, P. G. Platonov, Contribution of depolarization and repolarization changes to j-wave generation and ventricular fibrillation in ischemia, Front. physiol. 11 (2020). doi:10.3389/fphys.2020.568021.

[9] P. W. Macfarlane, C. Antzelevitch, M. Haissaguerre, H. V. Huikuri, M. Potse, R. Rosso, F. Sacher, J. T. Tikkanen, H. Wellens, G.-X. Yan, The early repolarization pattern, JACC 66 (4) (2015) 470–477. doi: 10.1016/j.jacc.2015.05.033.

