## Supplementary material for "Unmasking the J-wave: 3D ECG shows terminal depolarization mimicking early repolarization": Patient-level 3D ECG figures grouped by J-loop phenotype, showing planar leads and rotated 3D trajectories for all included cases.

### Supplementary 3D ECG figures by J-wave group

Alejandro Jesús Bermejo Valdés

#### Contents

Below is the list of all supplementary 3D ECG figures, organized by phenotype group. Click on any entry to jump to the corresponding figure.

##### Group i


- Figure: patient 017 (s0053lre)
- Figure: patient 040 (s0133lre)
- Figure: patient 045 (s0147lre)
- Figure: patient 048 (s0171lre)
- Figure: patient 049 (s0173lre)
- Figure: patient 052 (s0190lre)
- Figure: patient 057 (s0198lre)
- Figure: patient 083 (s0268lre)
- Figure: patient 099 (s0387lre)
- Figure: patient 110 (s0003_re)
- Figure: patient 165 (s0322lre)
- Figure: patient 173 (s0305lre)
- Figure: patient 180 (s0374lre)
- Figure: patient 198 (s0402lre)
- Figure: patient 240 (s0468_re)
- Figure: patient 241 (s0469_re)
- Figure: patient 245 (s0474_re)
- Figure: patient 248 (s0481_re)
- Figure: patient 249 (s0484_re)
- Figure: patient 250 (s0485_re)
- Figure: patient 252 (s0487_re)
- Figure: patient 258 (s0494_re)

- Figure: patient 259 (s0495_re)
- Figure: patient 273 (s0511_re)
- Figure: patient 277 (s0527_re)

##### Group ii

- Figure: patient 043 (s0141lre)
- Figure: patient 230 (s0454_re)
- Figure: patient 244 (s0473_re)

##### Group iii

- Figure: patient 073 (s0238lre)
- Figure: patient 150 (s0287lre)
- Figure: patient 187 (s0207_re)

#### Supplementary Figures

All figures share the same layout. The top row displays leads V1-V6. The middle and bottom rows show the 3D trajectory in the (V1, V6, time) space from different viewing angles. The J-related segment of the terminal QRS loop is highlighted in red.

#### Group i

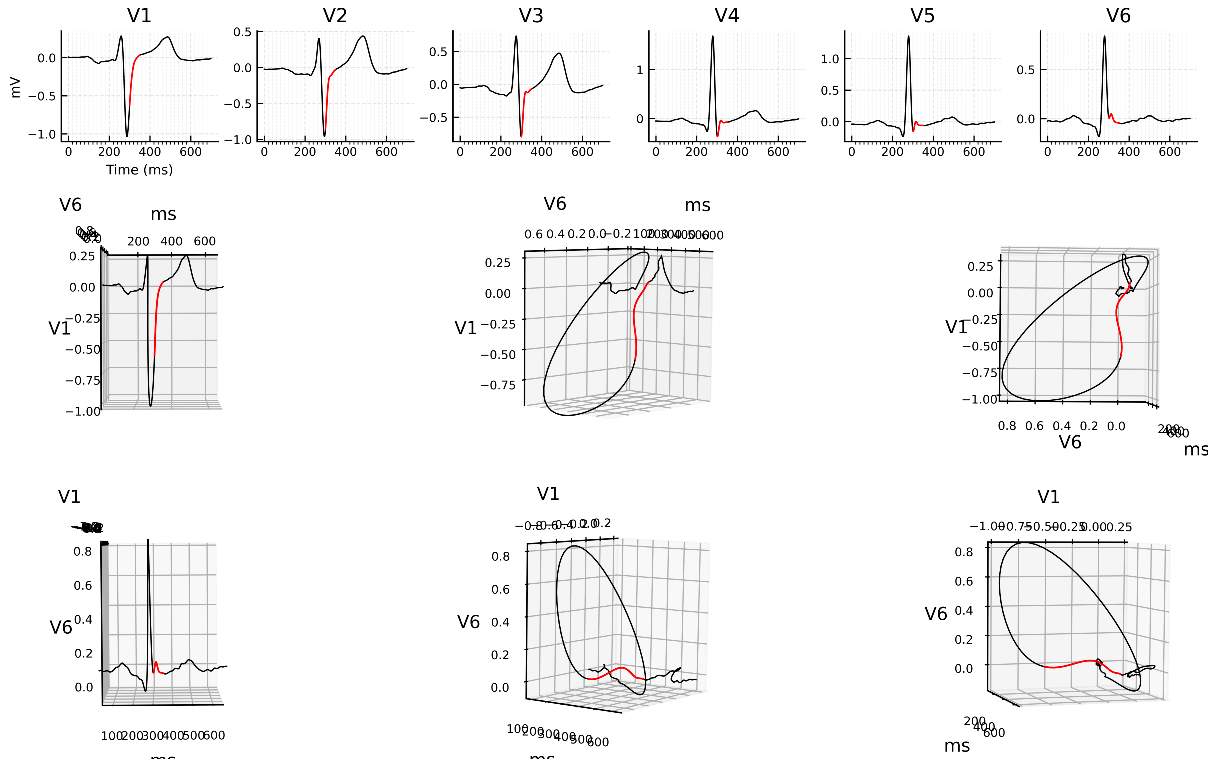


Figure 1: 3D ECG example for patient 017 (record s0053lre), group i.

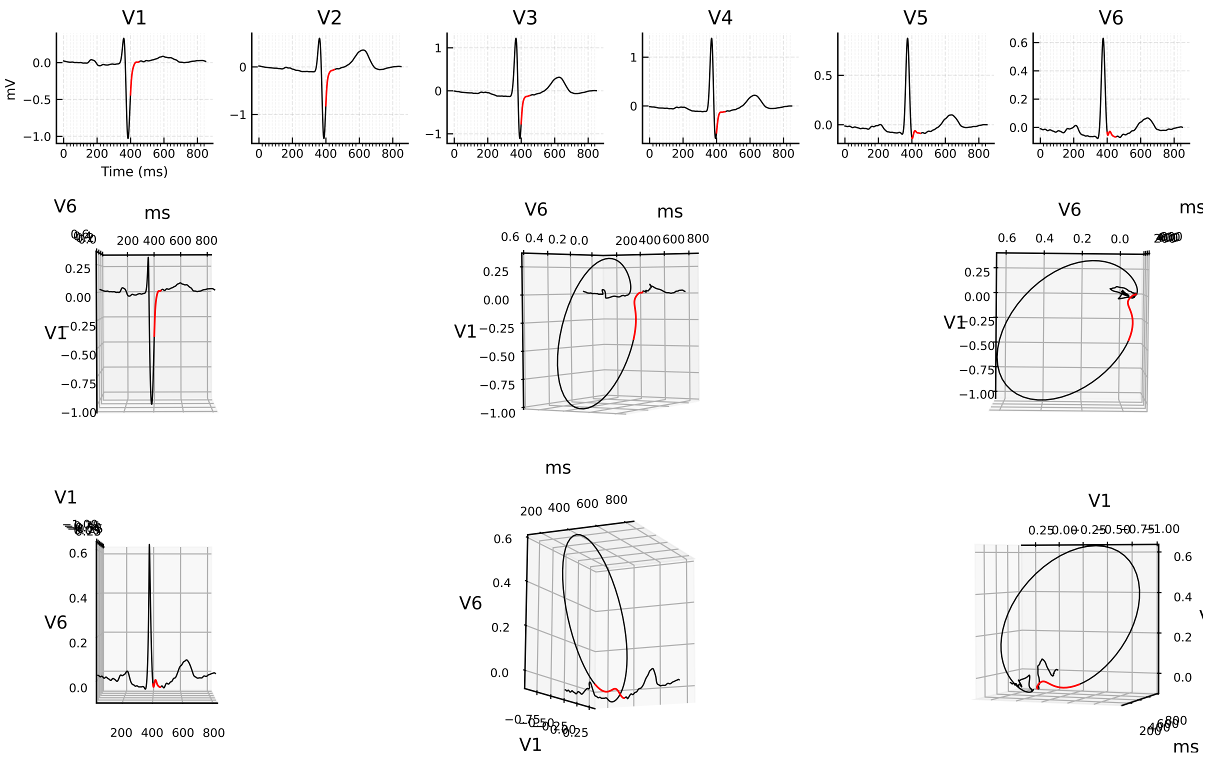


Figure 2: 3D ECG example for patient 040 (record s0133lre), group i.

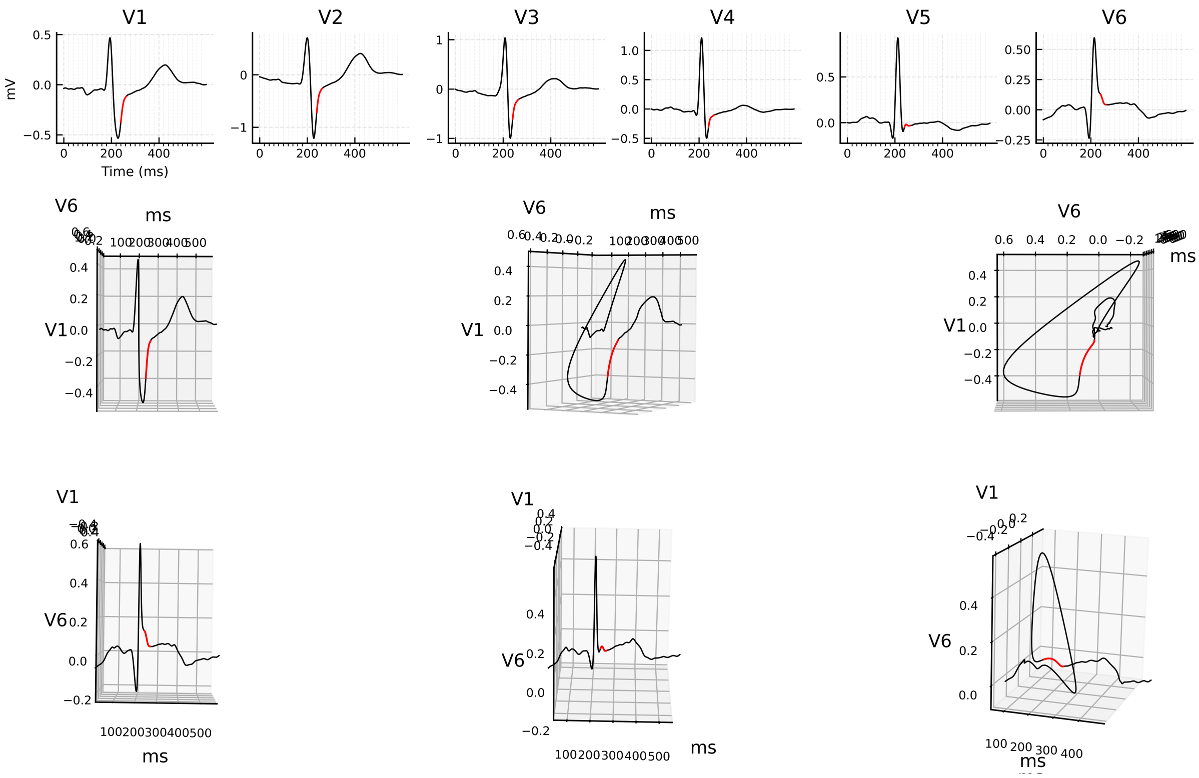


Figure 3: 3D ECG example for patient 045 (record s0147lre), group i.

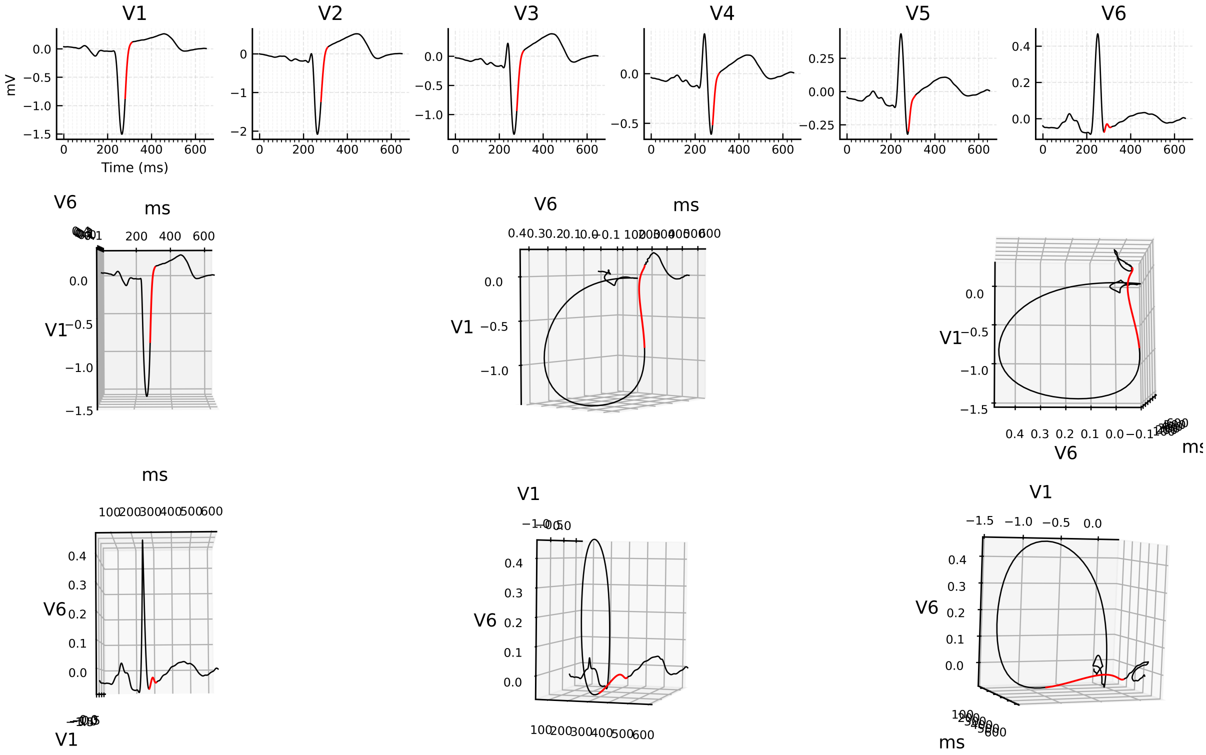


Figure 4: 3D ECG example for patient 048 (record s0171lre), group i.

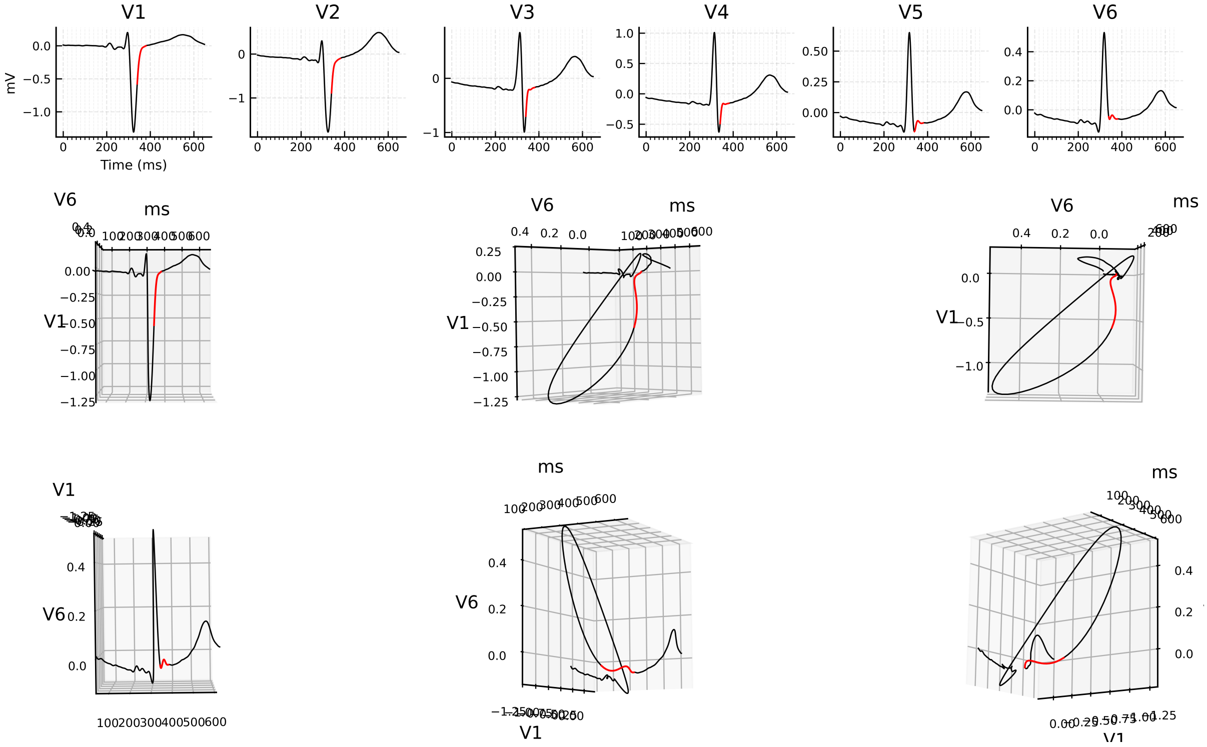


Figure 5: 3D ECG example for patient 049 (record s0173lre), group i.

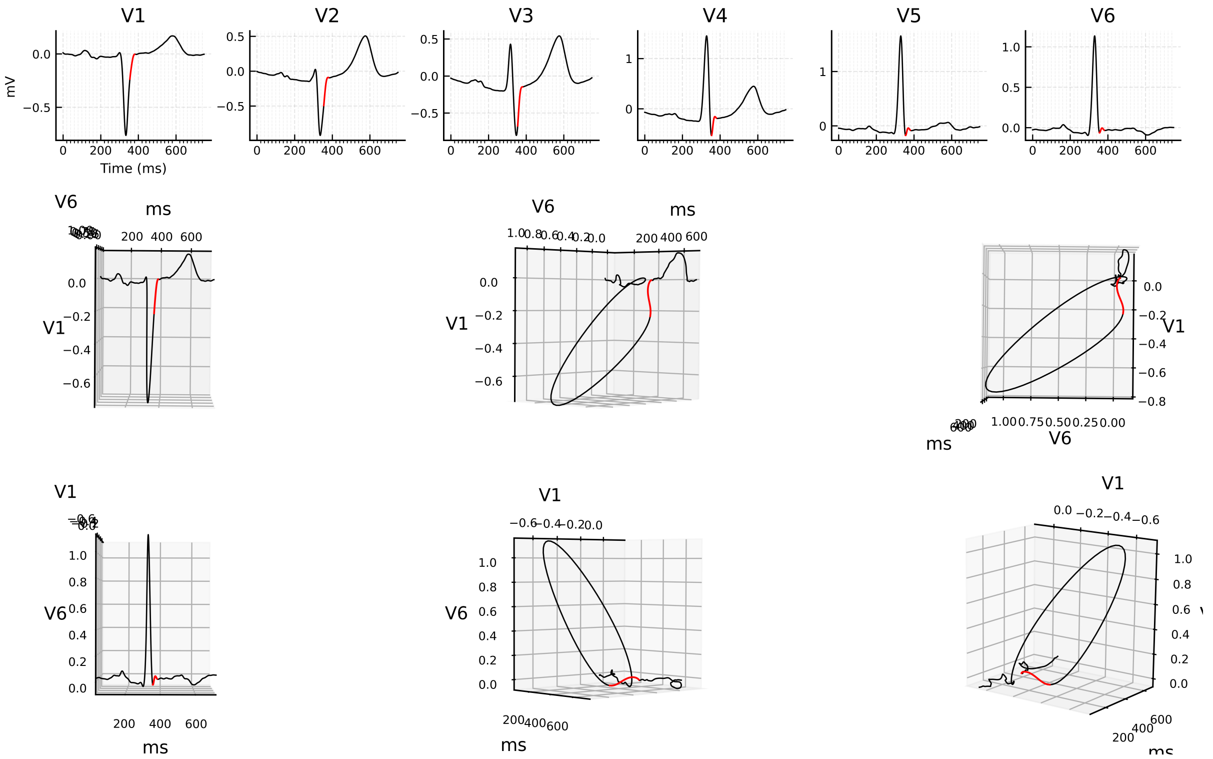


Figure 6: 3D ECG example for patient 052 (record s0190lre), group i.

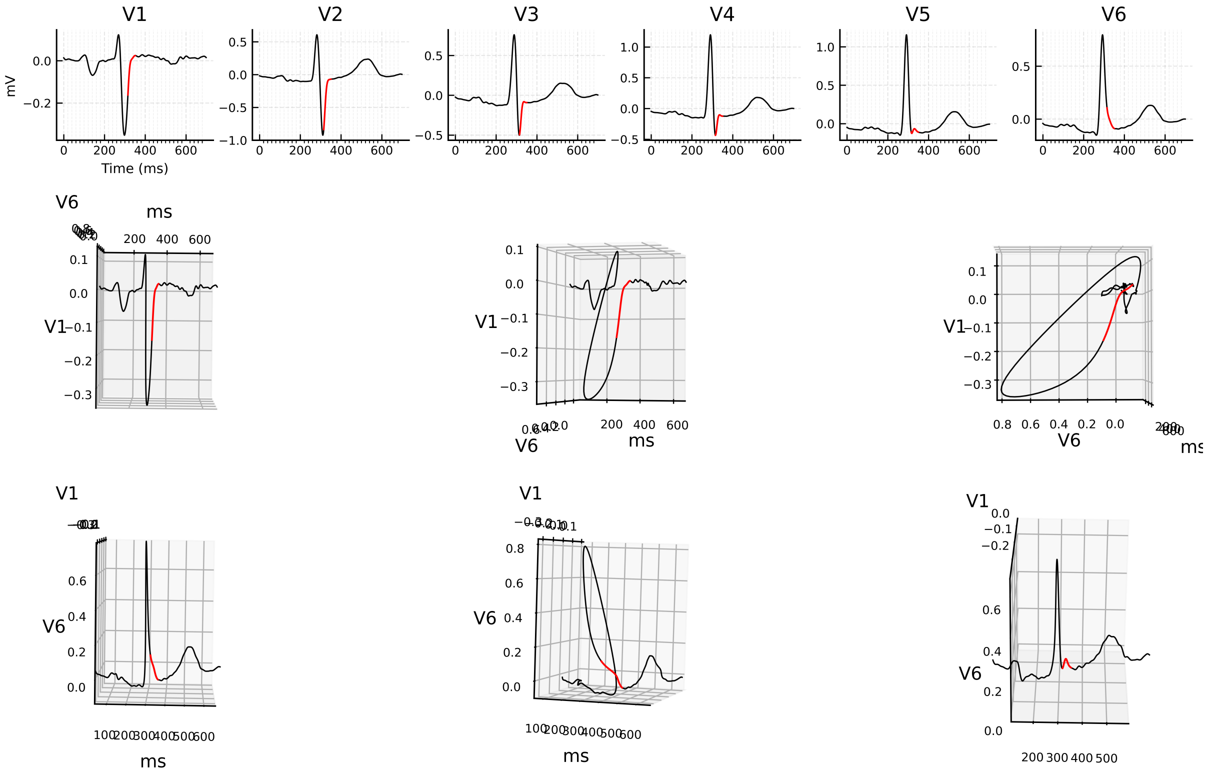


Figure 7: 3D ECG example for patient 057 (record s0198lre), group i.

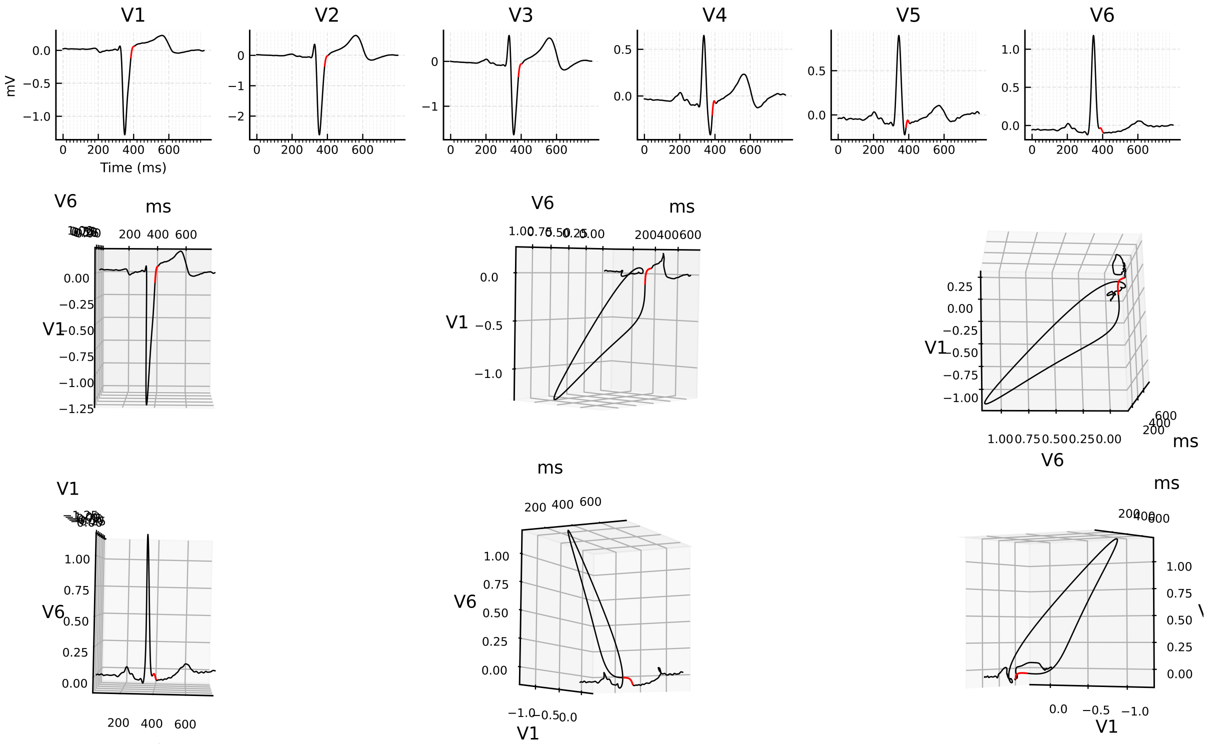


Figure 8: 3D ECG example for patient 083 (record s0268lre), group i.

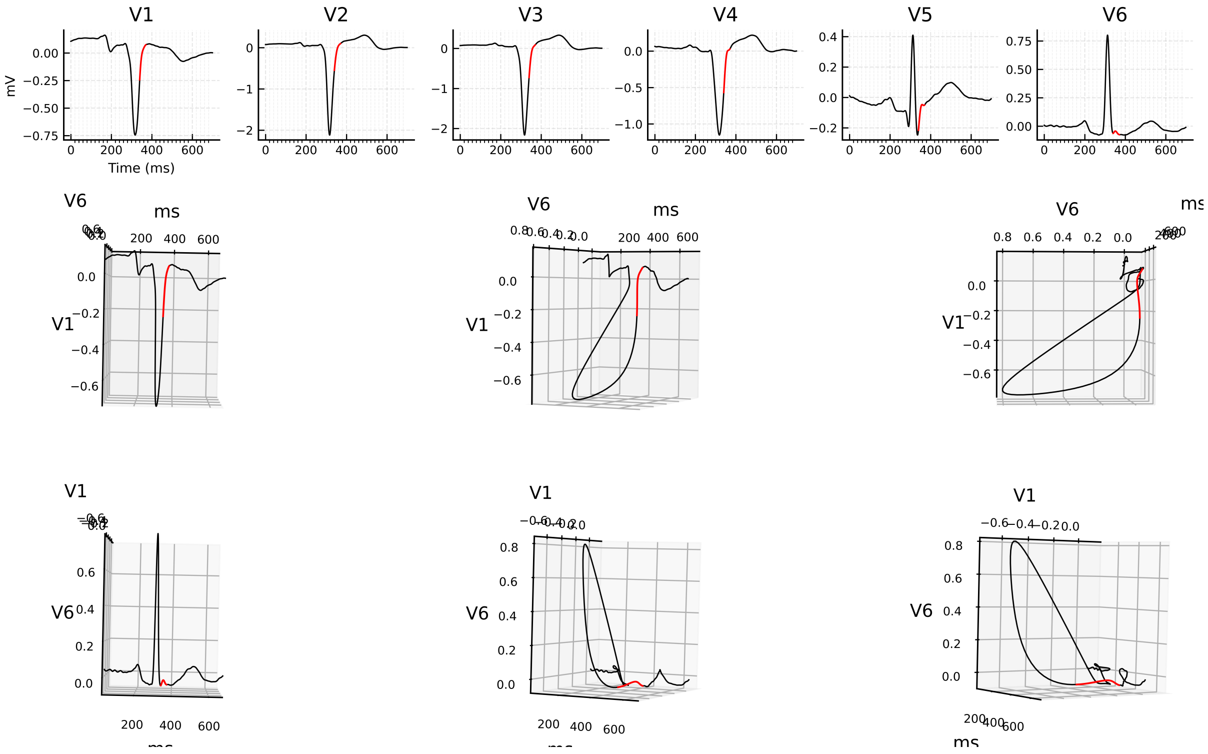


Figure 9: 3D ECG example for patient 099 (record s0387lre), group i.

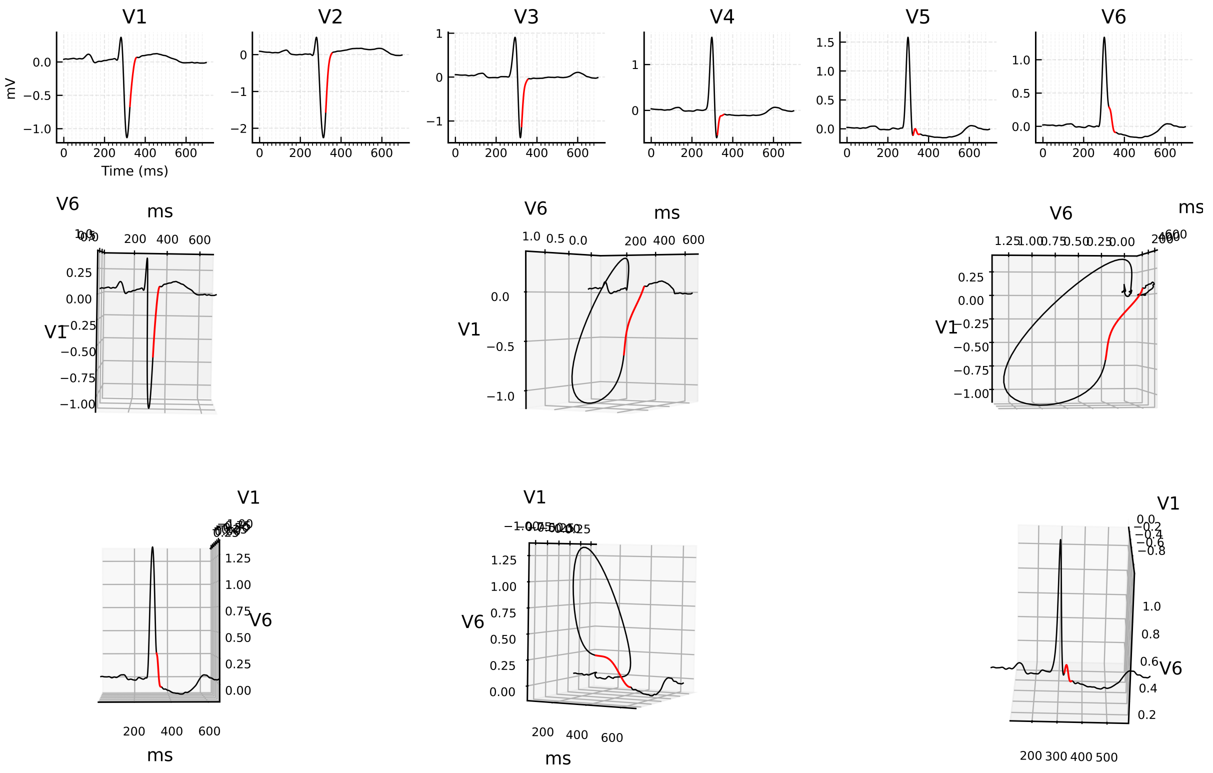


Figure 10: 3D ECG example for patient 110 (record s0003_re), group i.

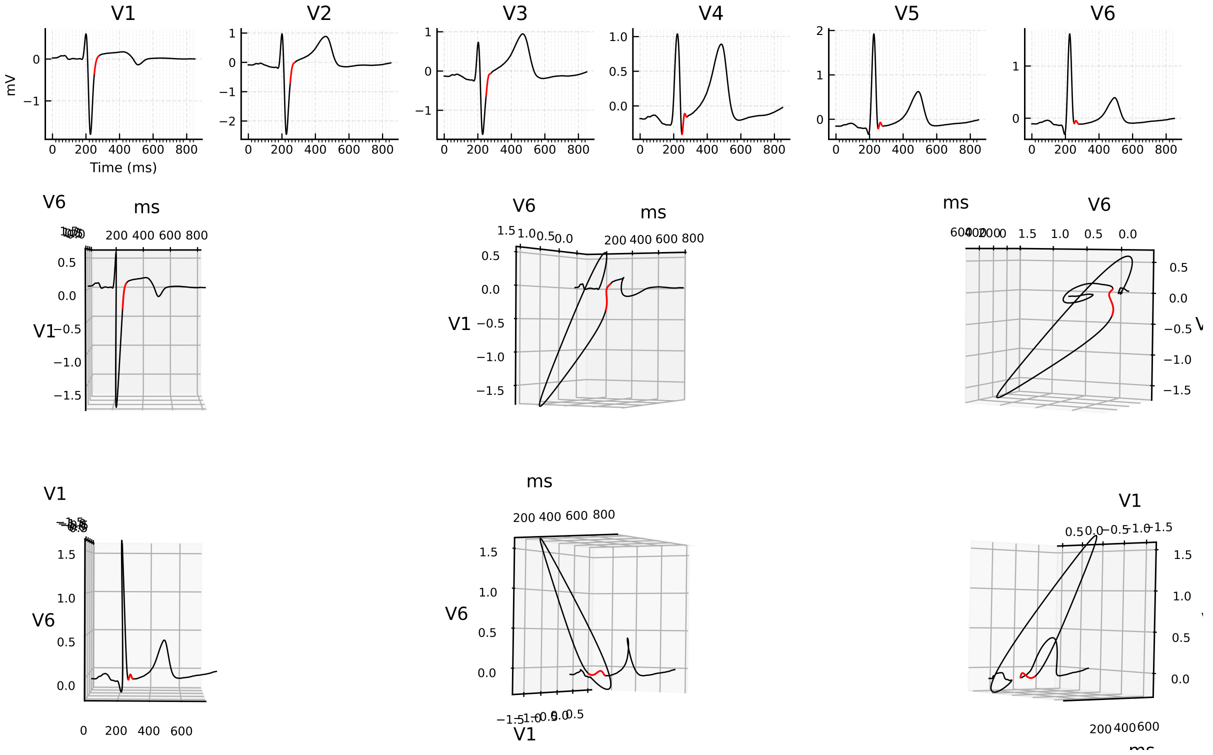


Figure 11: 3D ECG example for patient 165 (record s0322lre), group i.

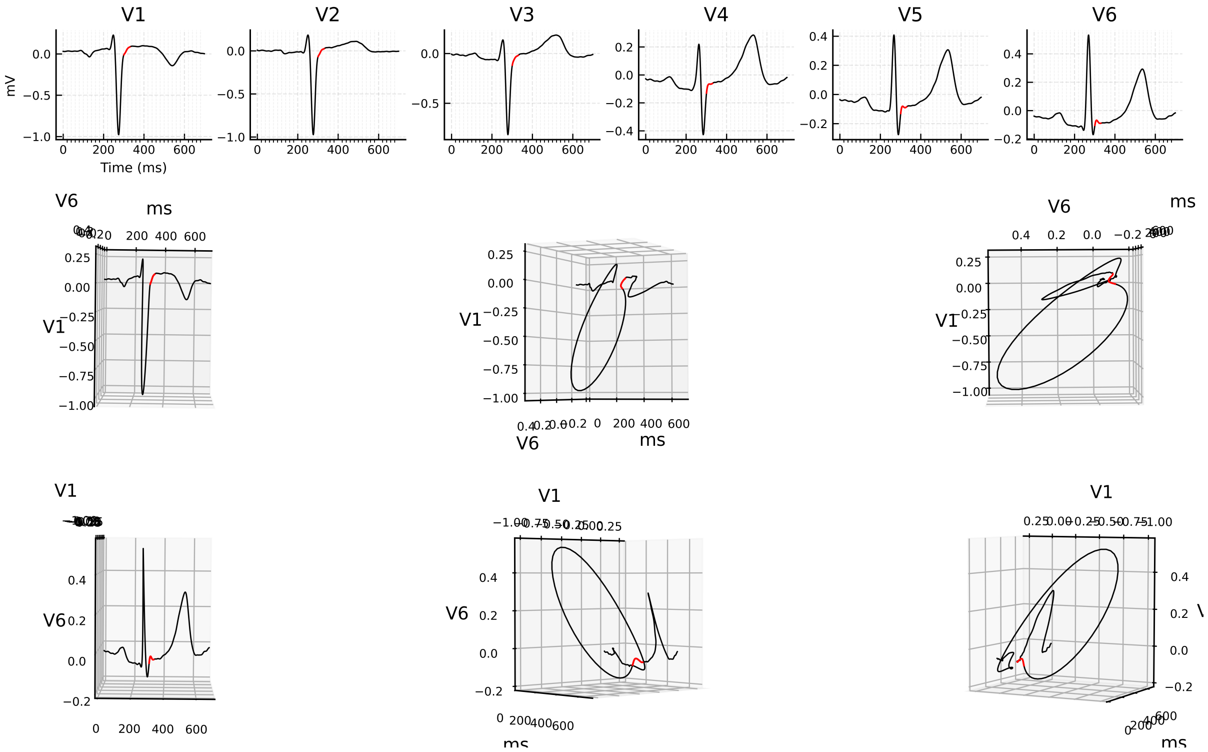


Figure 12: 3D ECG example for patient 173 (record s0305lre), group i.

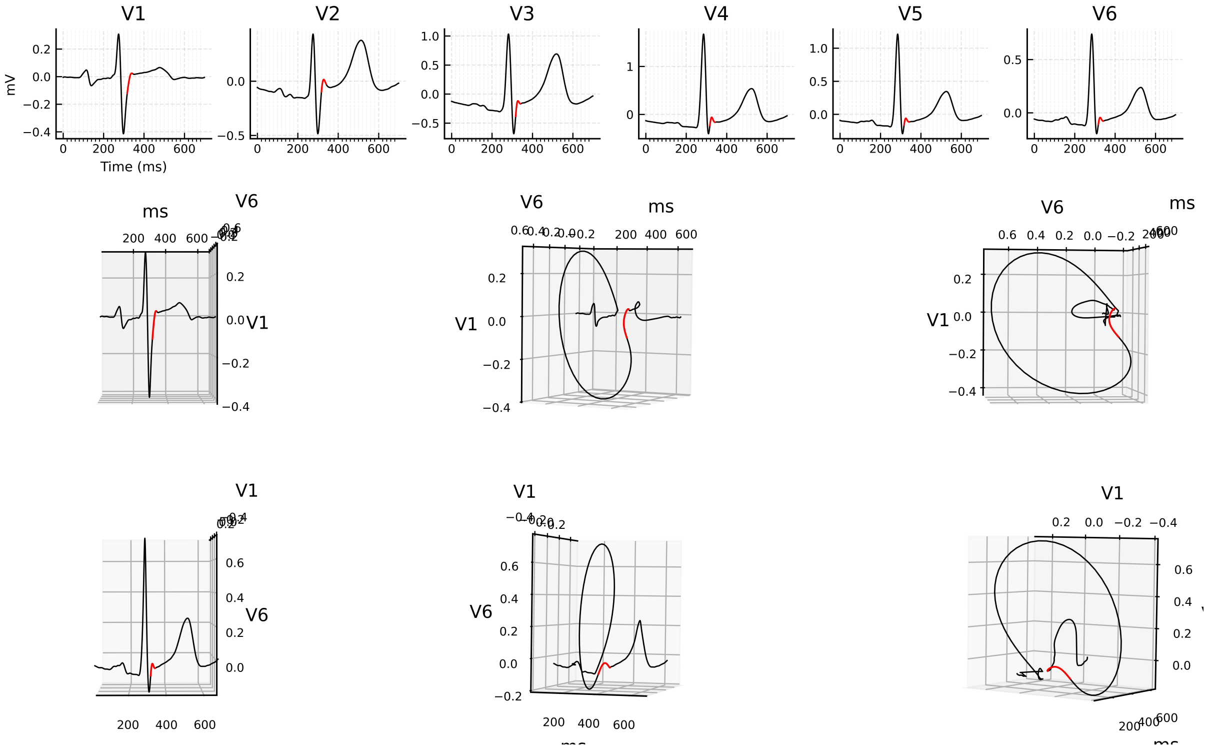


Figure 13: 3D ECG example for patient 180 (record s0374lre), group i.

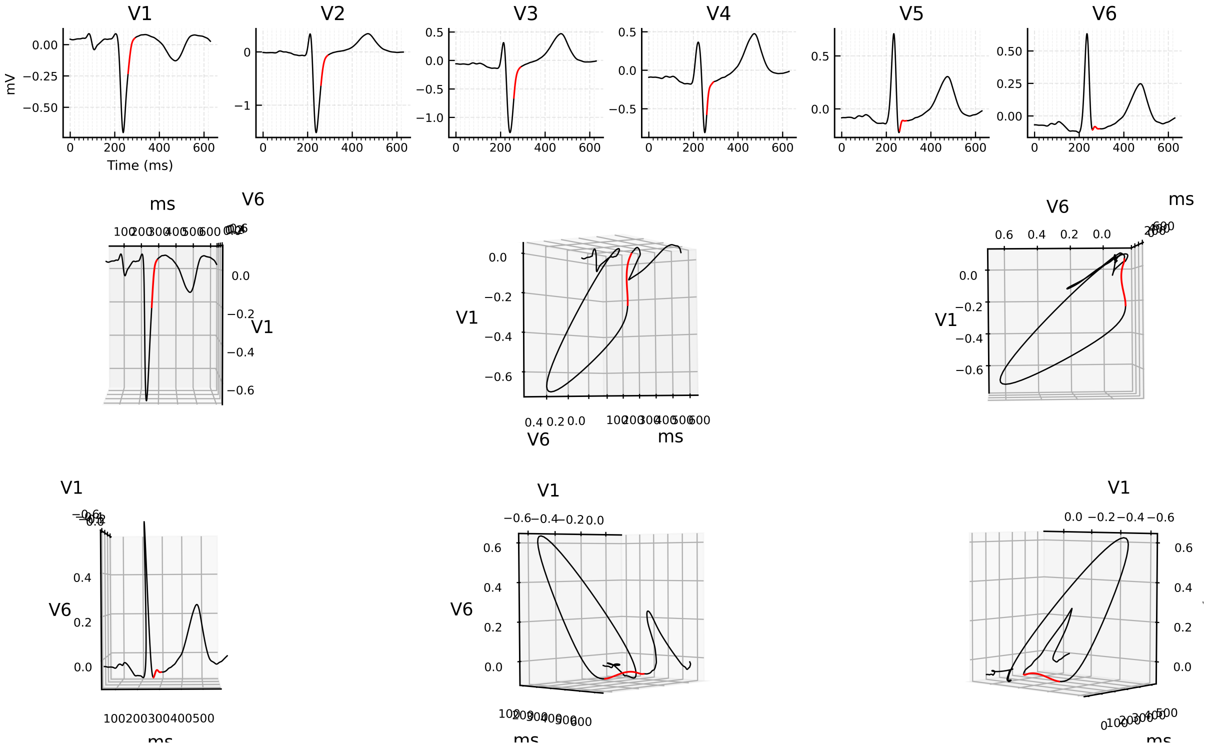


Figure 14: 3D ECG example for patient 198 (record s0402lre), group i.

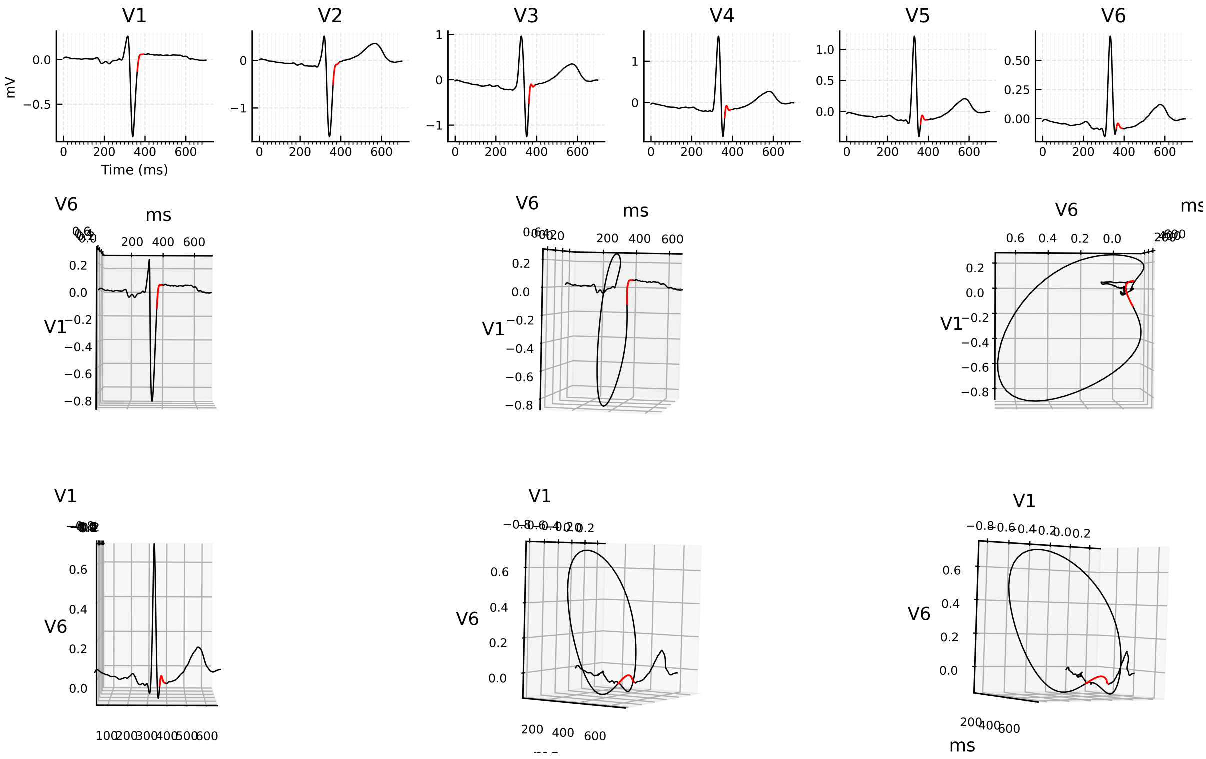


Figure 15: 3D ECG example for patient 240 (record s0468_re), group i.

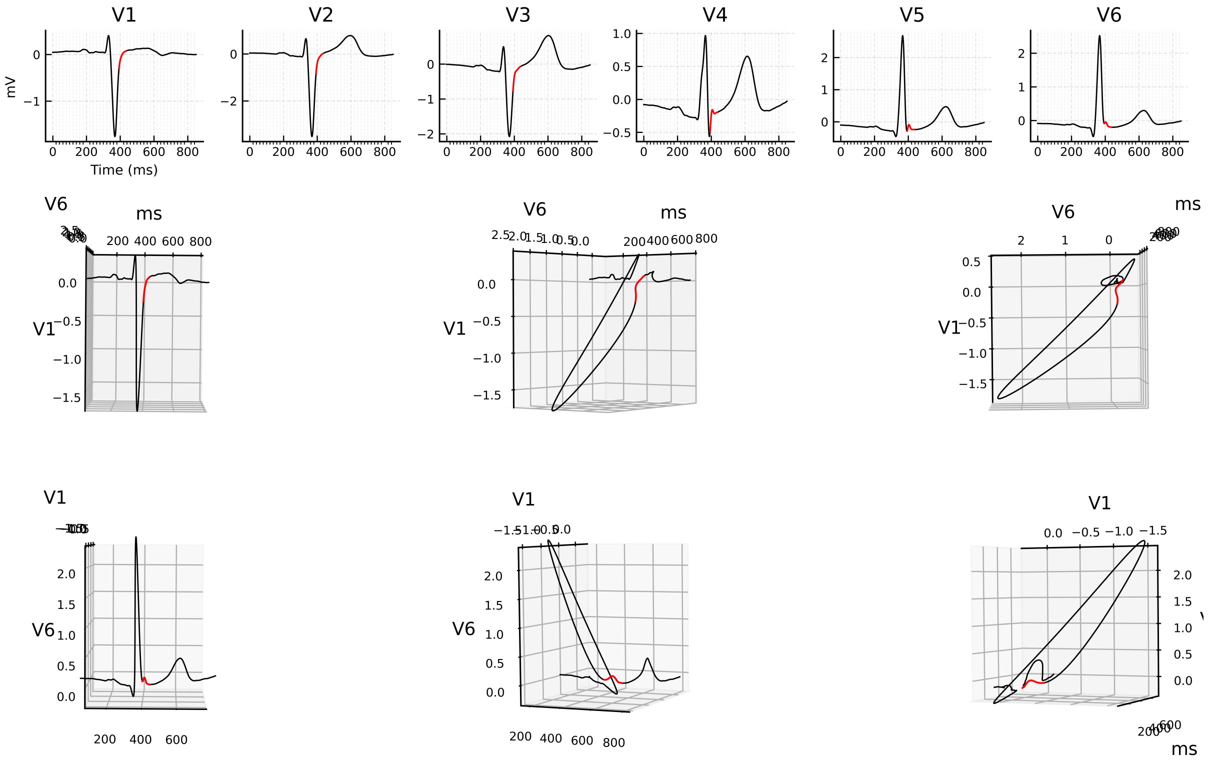

Figure 16: 3D ECG example for patient 241 (record s0469_re), group i.

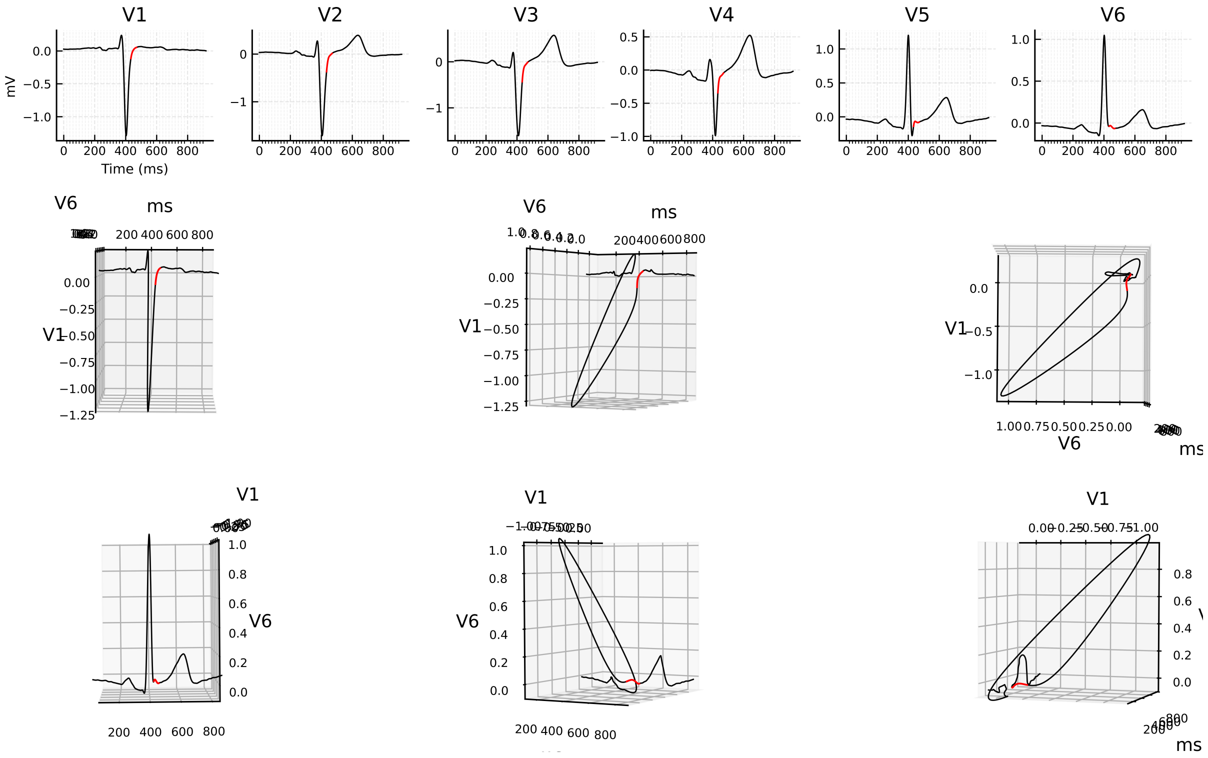

Figure 17: 3D ECG example for patient 245 (record s0474_re), group i.

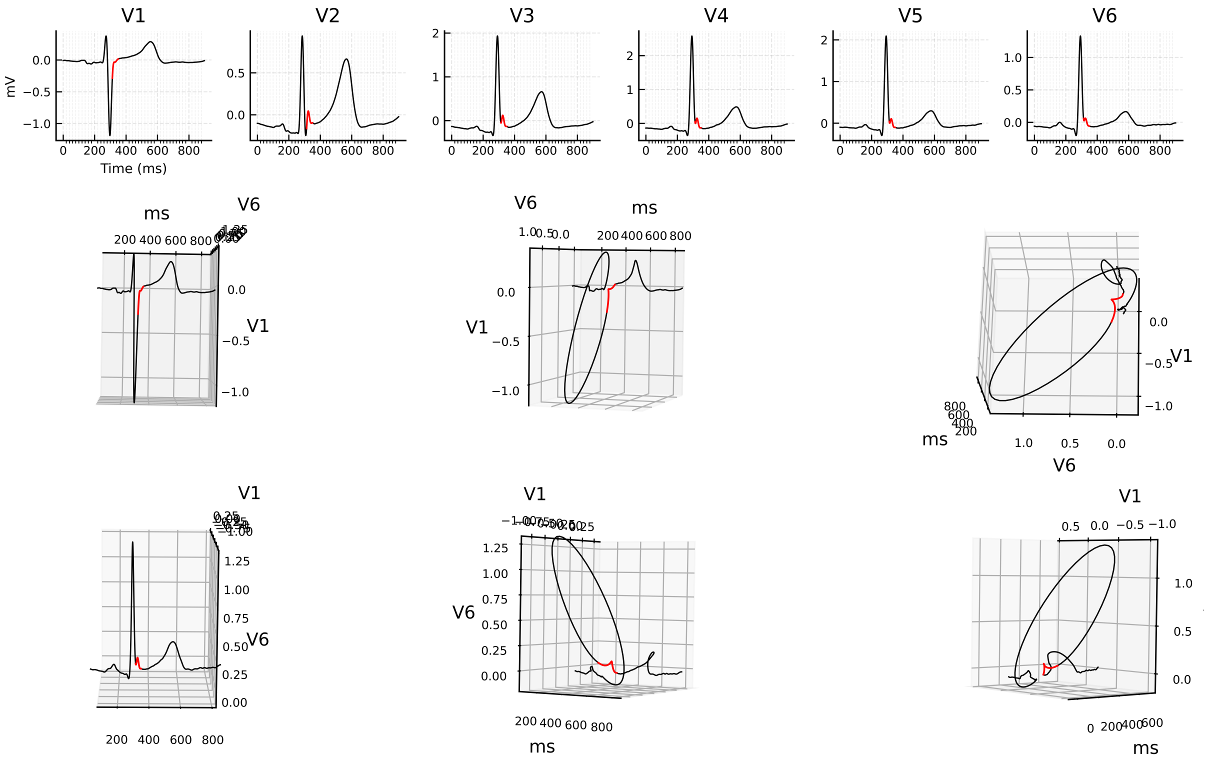


Figure 18: 3D ECG example for patient 248 (record s0481_re), group i.

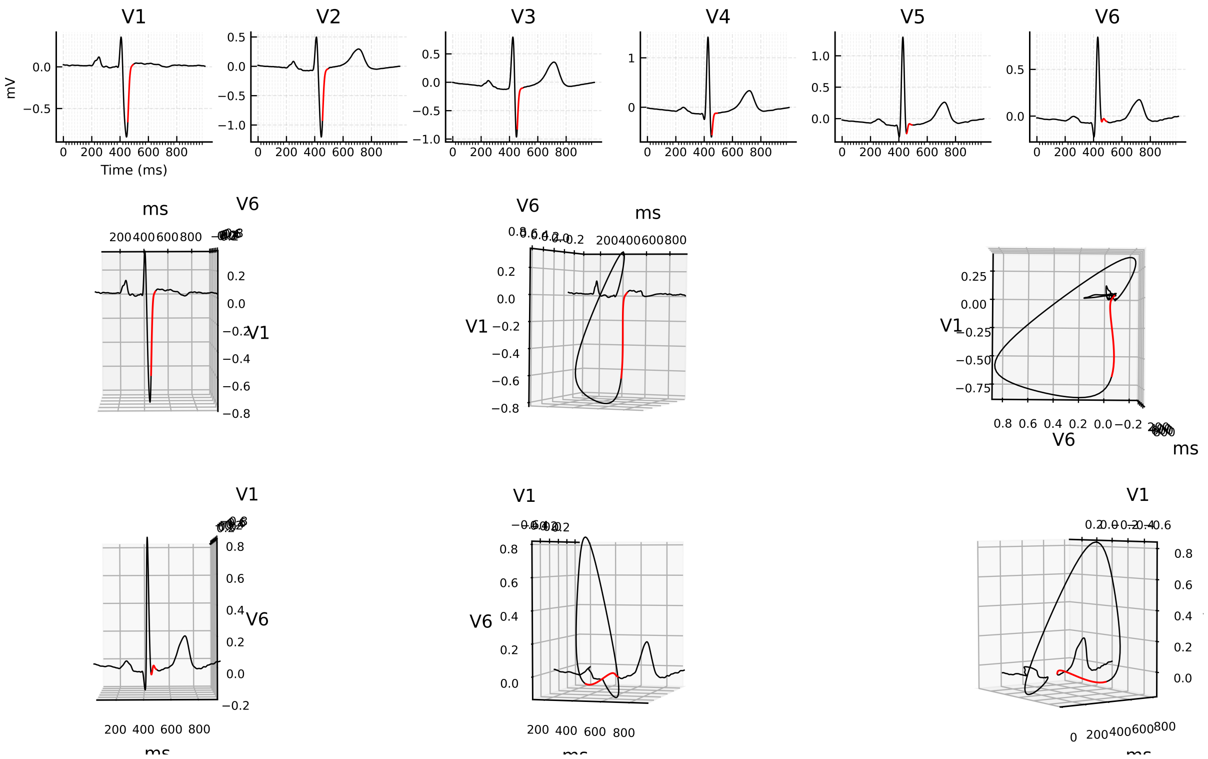


Figure 19: 3D ECG example for patient 249 (record s0484_re), group i.

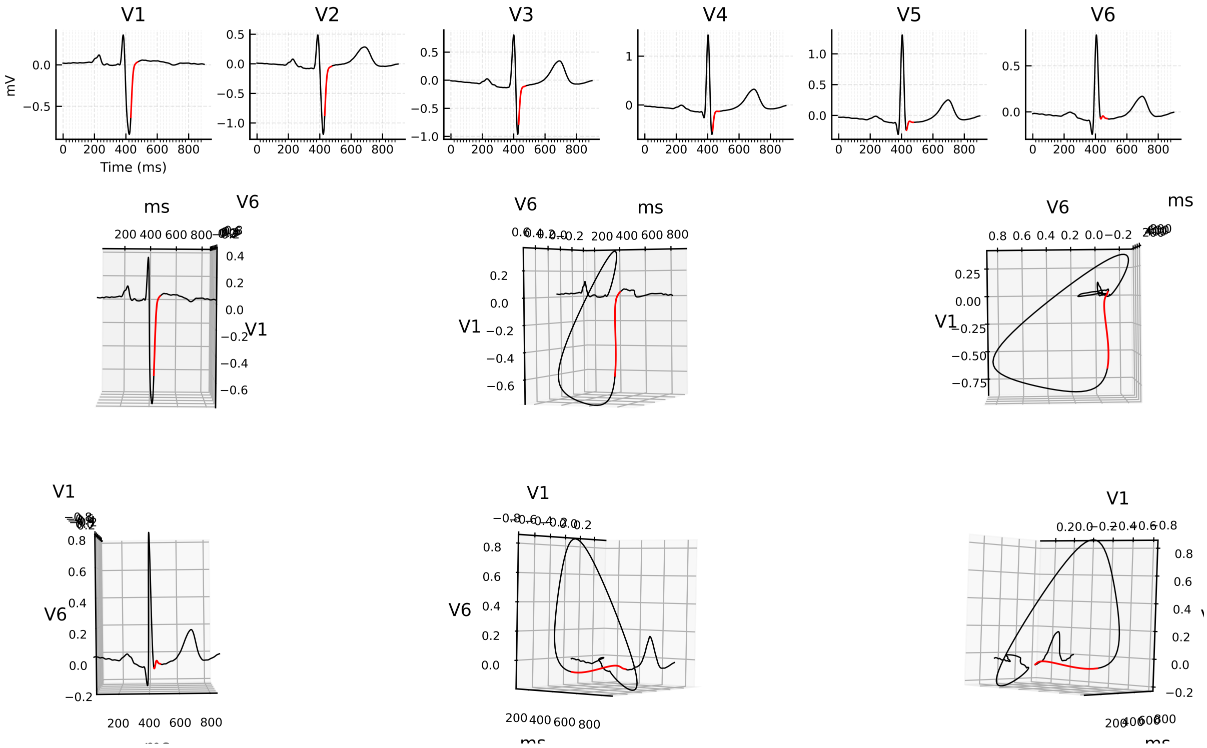


Figure 20: 3D ECG example for patient 250 (record s0485_re), group i.

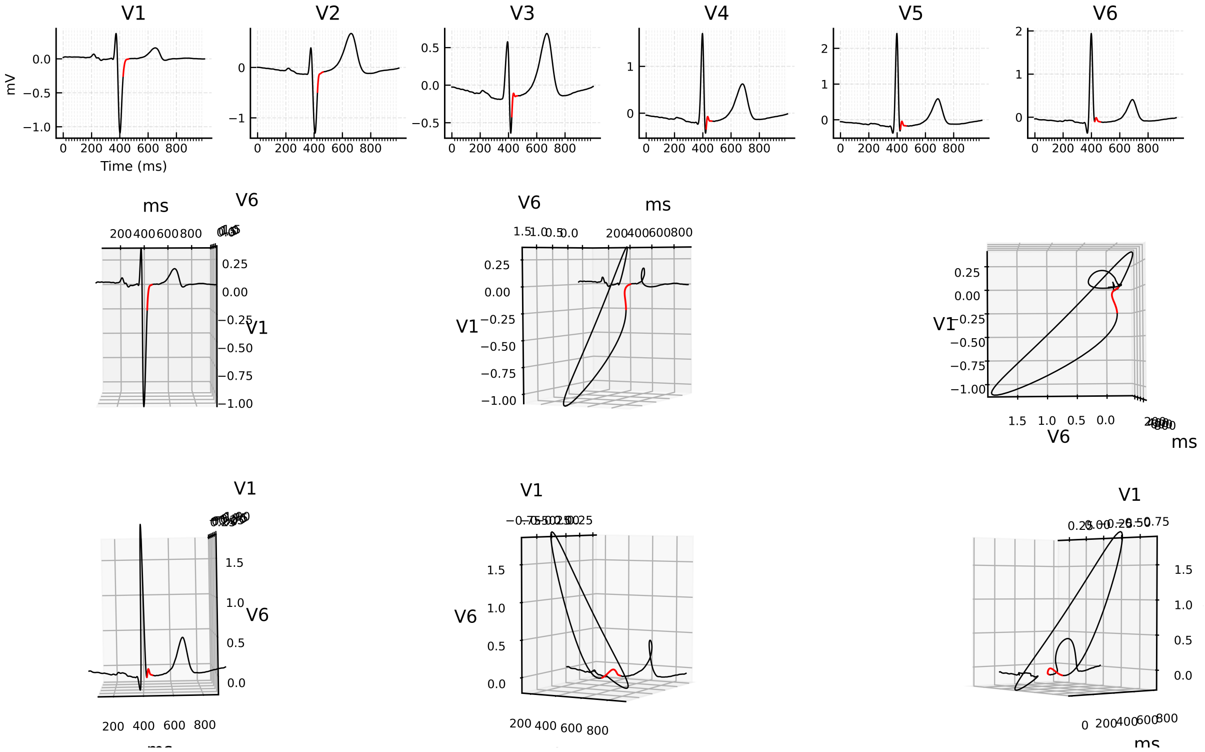


Figure 21: 3D ECG example for patient 252 (record s0487_re), group i.

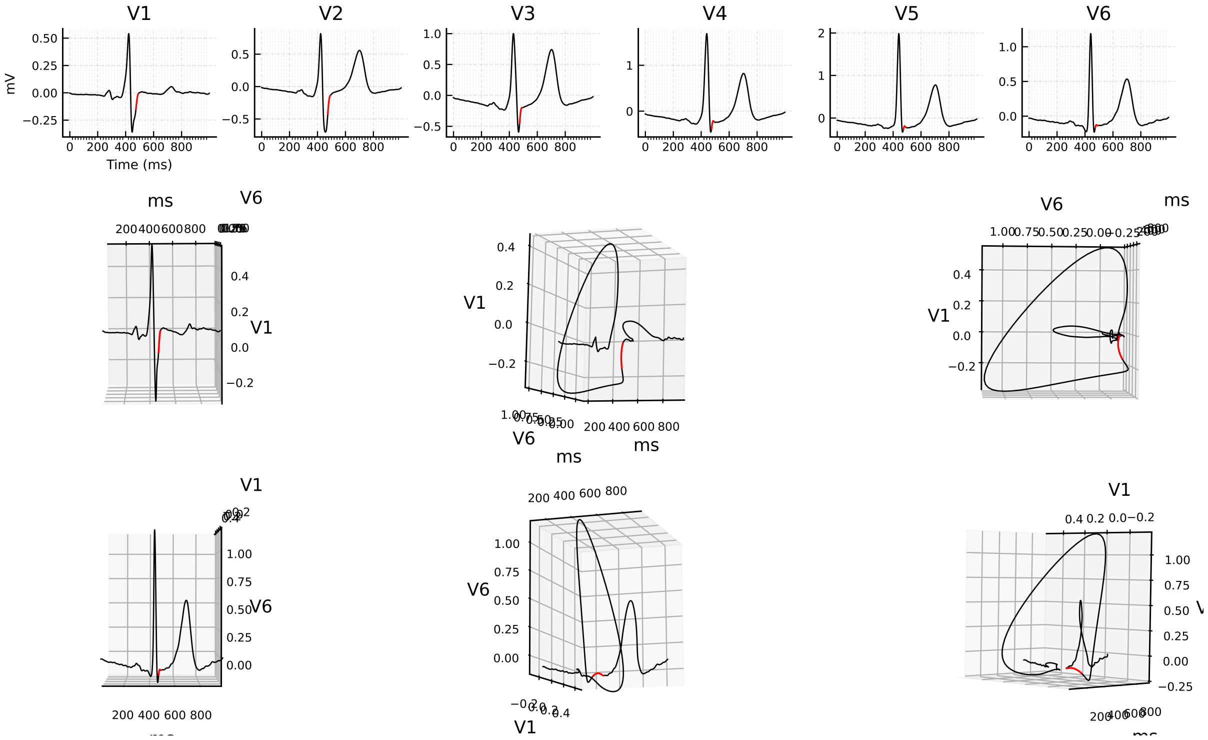

Figure 22: 3D ECG example for patient 258 (record s0494_re), group i.

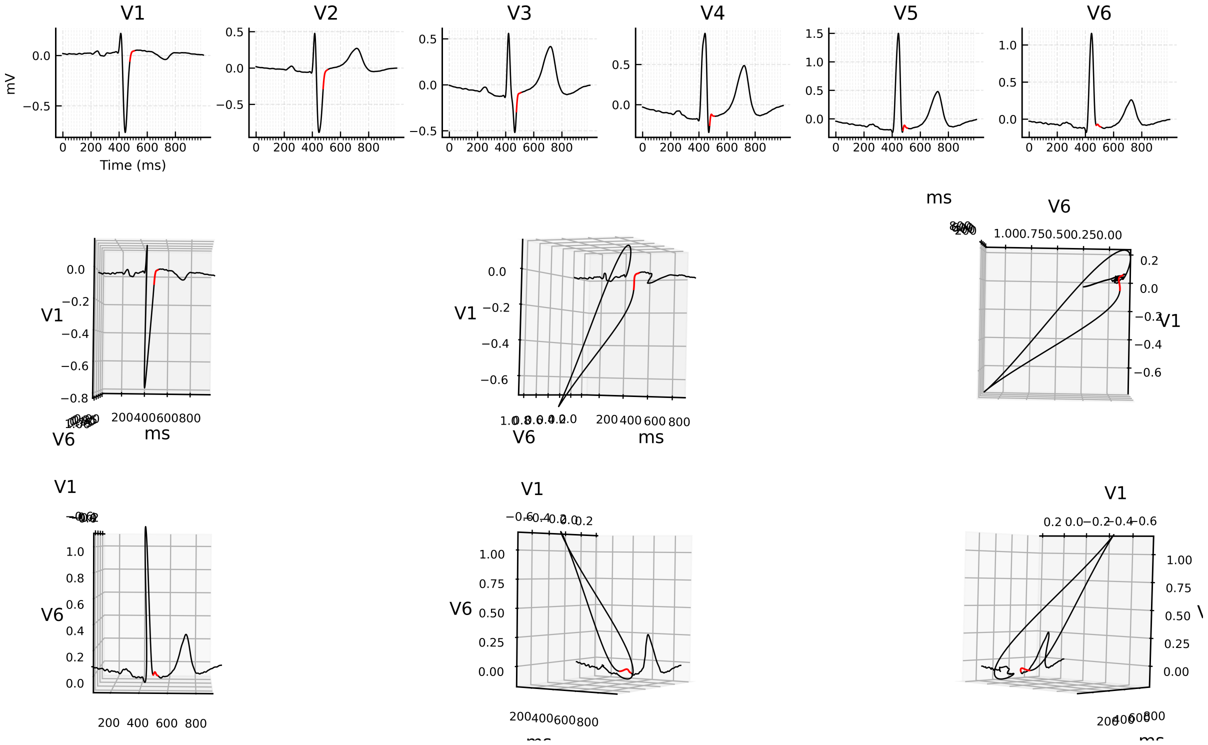

Figure 23: 3D ECG example for patient 259 (record s0495_re), group i.

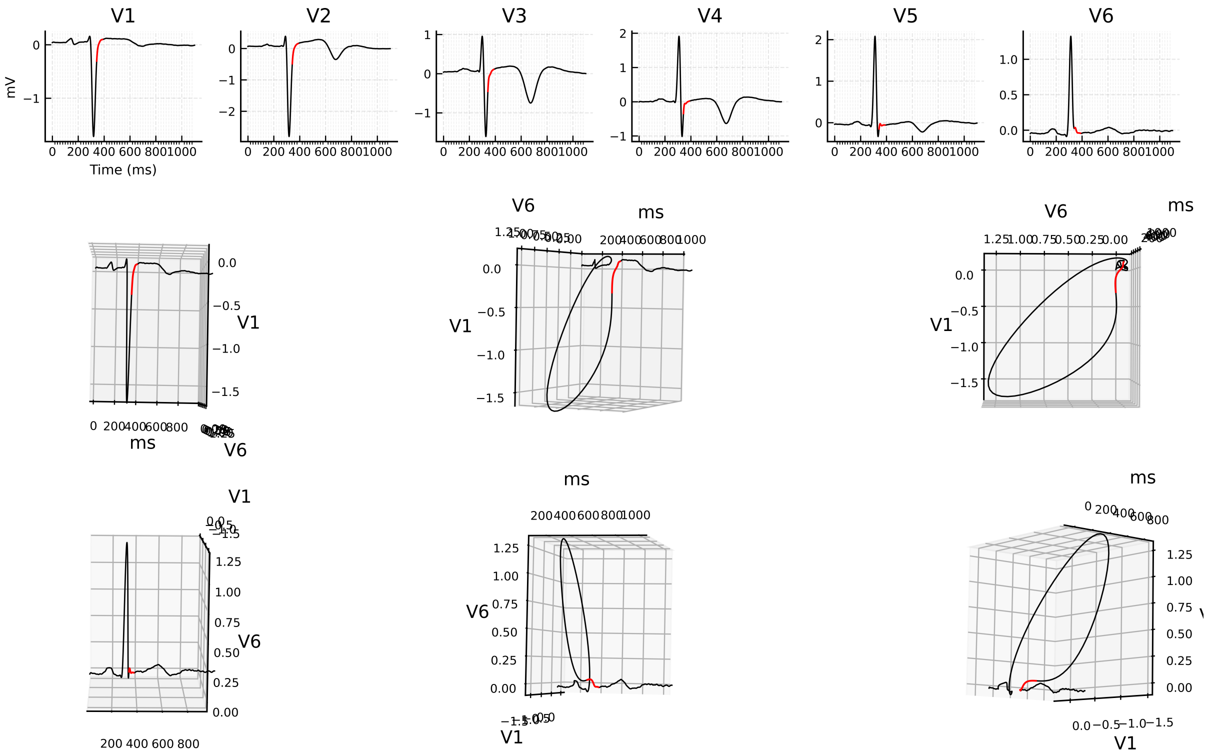


Figure 24: 3D ECG example for patient 273 (record s0511_re), group i.

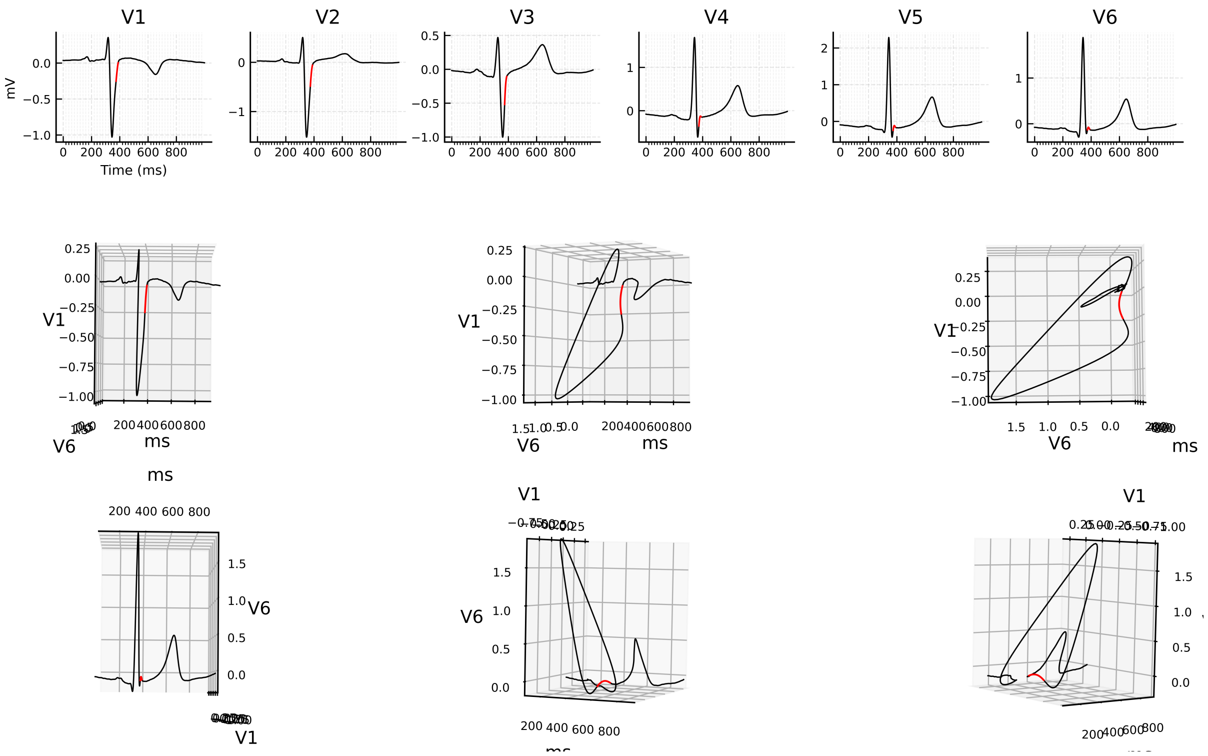


Figure 25: 3D ECG example for patient 277 (record s0527_re), group i.

#### Group ii

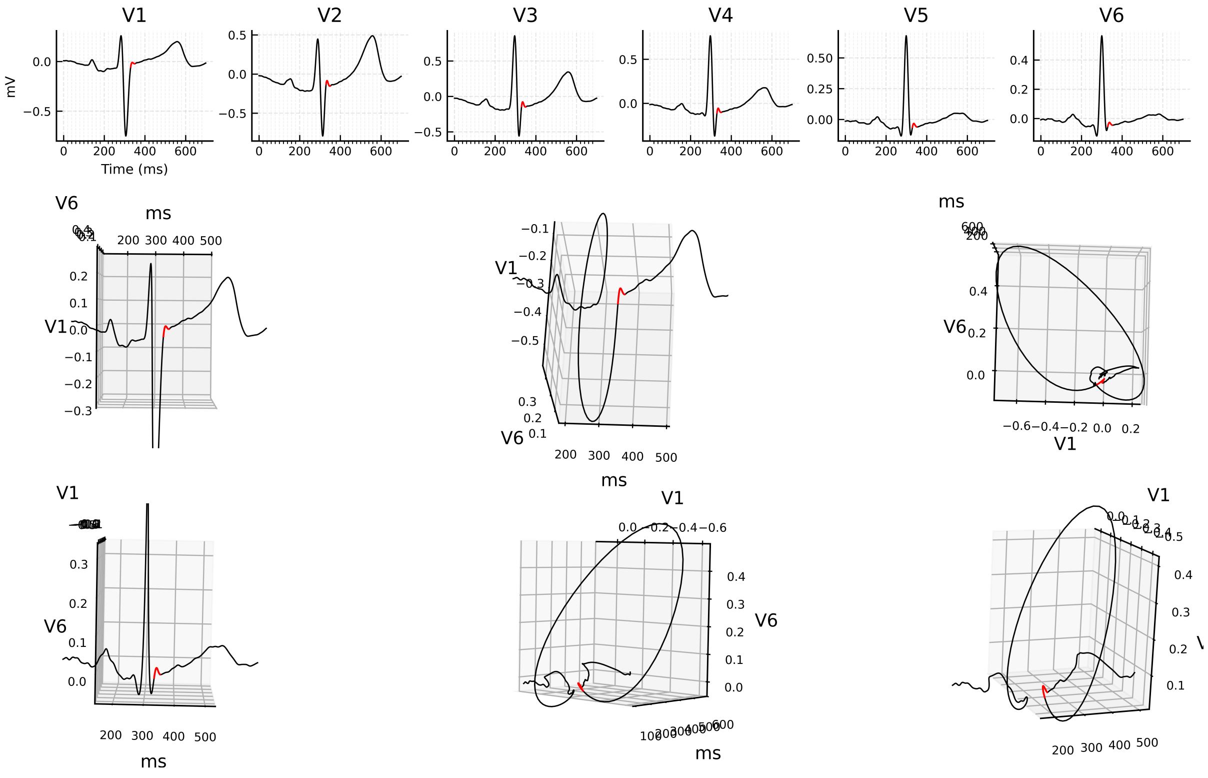


Figure 26: 3D ECG example for patient 043 (record s0141lre), group ii.

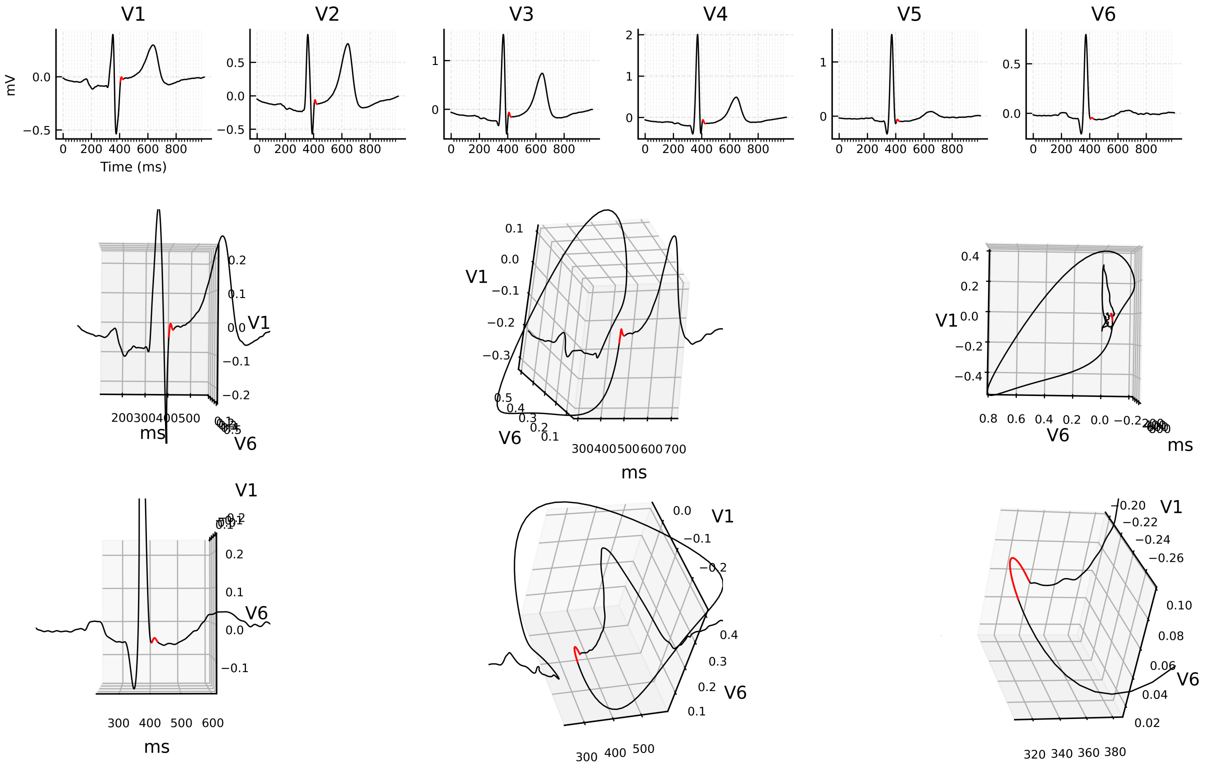


Figure 27: 3D ECG example for patient 230 (record s0454_re), group ii.

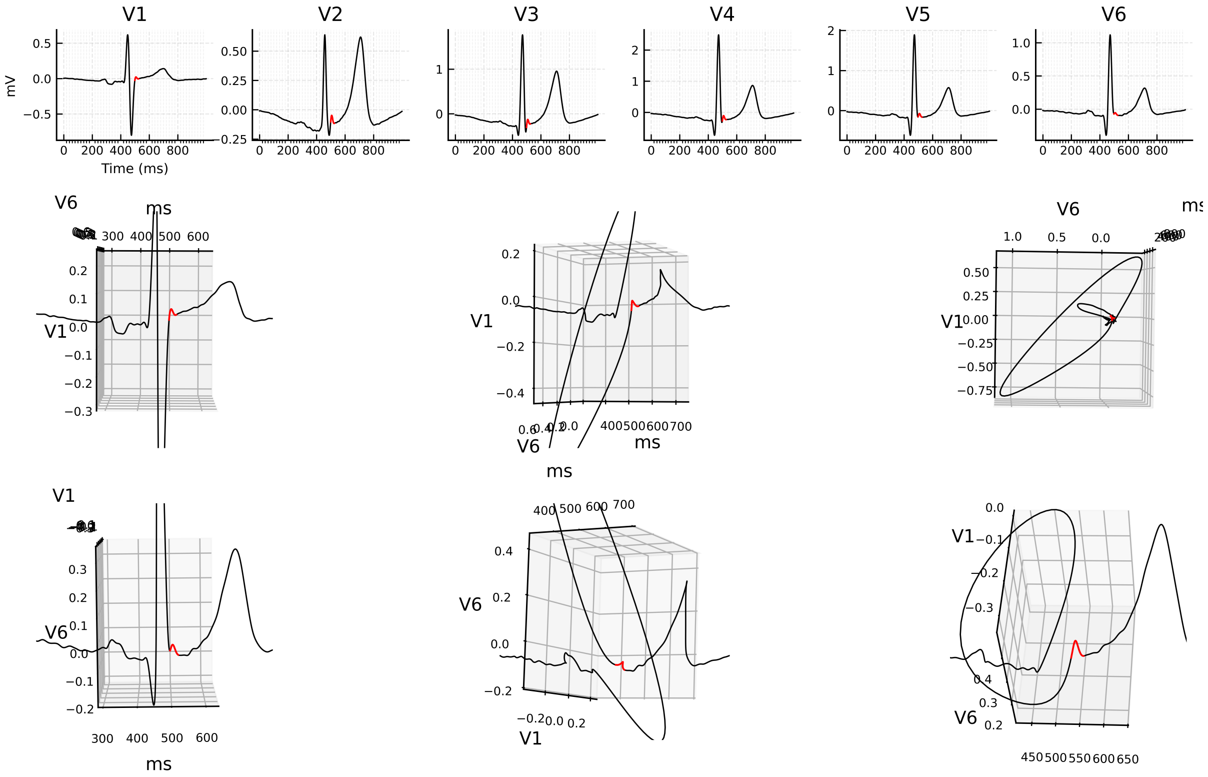


Figure 28: 3D ECG example for patient 244 (record s0473_re), group ii.

#### Group iii

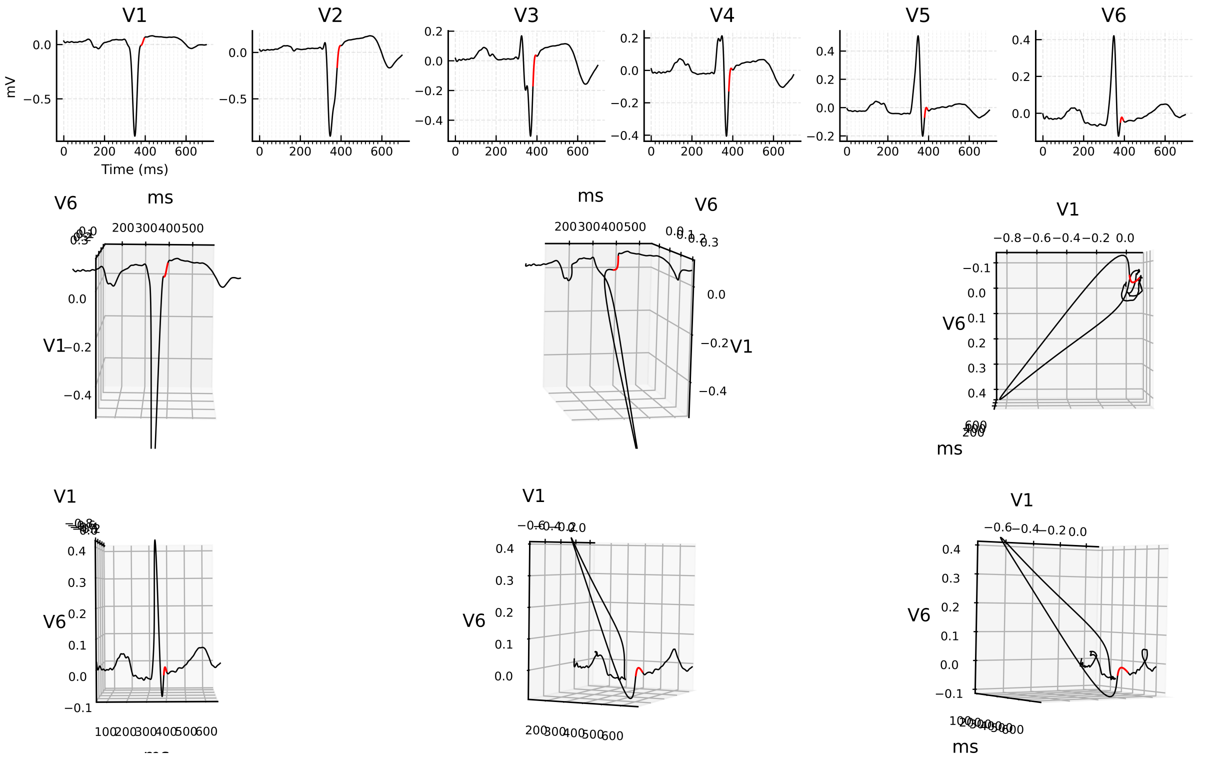


Figure 29: 3D ECG example for patient 073 (record s0238lre), group iii.

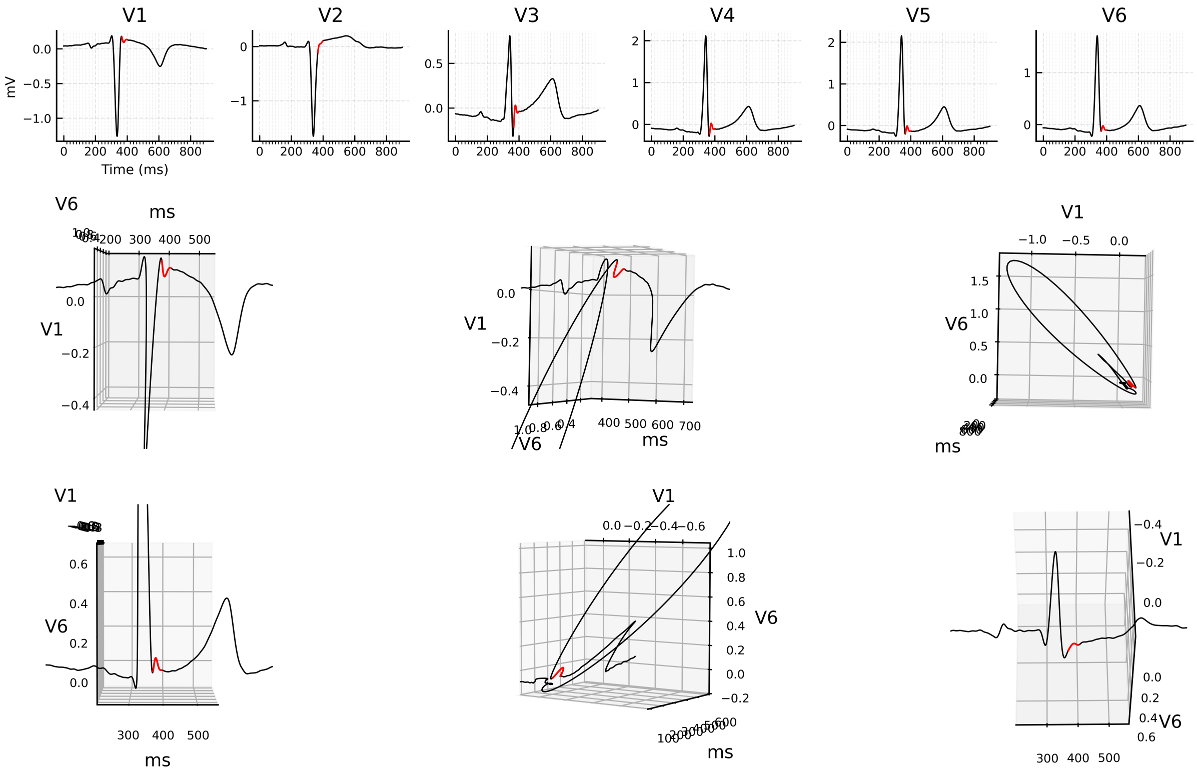


Figure 30: 3D ECG example for patient 150 (record s0287lre), group iii.

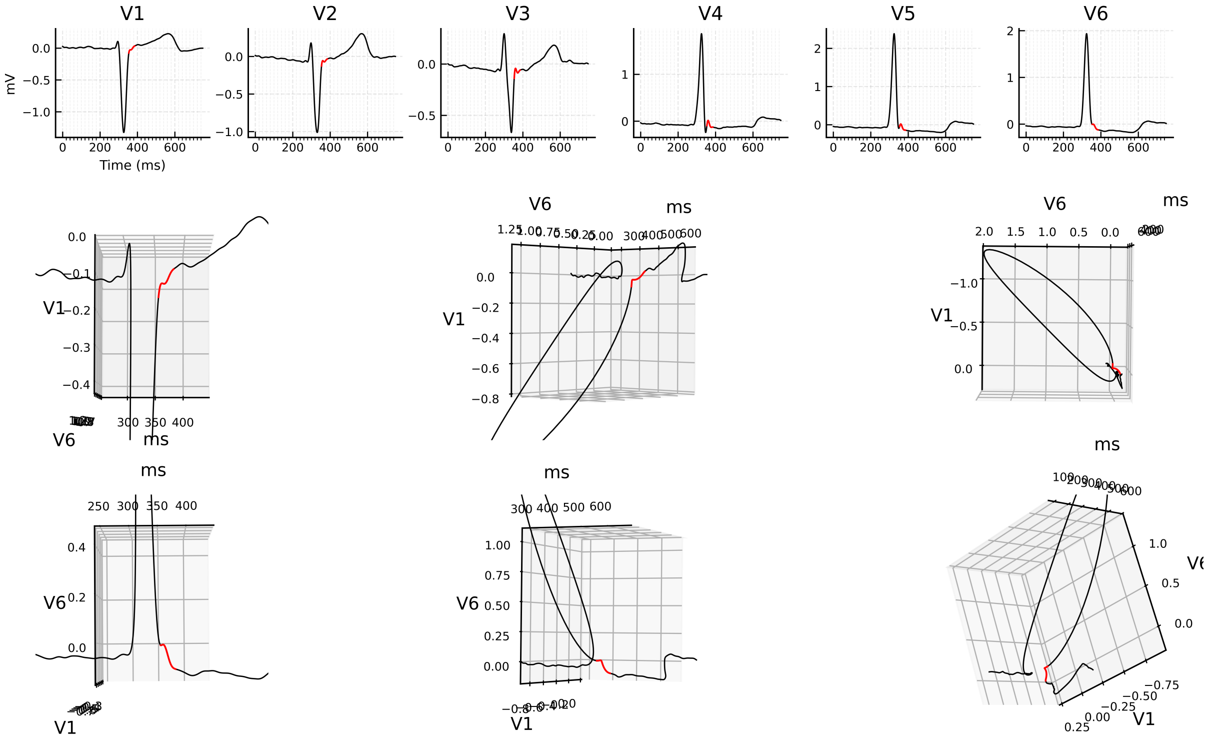


Figure 31: 3D ECG example for patient 187 (record s0207_re), group iii.
