## Supplementary material for "Unmasking the J-wave: 3D ECG shows terminal depolarization mimicking early repolarization": Patient-by-patient classification of V6 terminal QRS deflections and the corresponding rotational transformations, including excluded cases.

**Supplementary classification of J-wave and slurring phenotypes**

Alejandro J. Bermejo Valdés

This supplementary document summarizes all cases ($N = 31$) according to their V6 terminal QRS morphology (J-wave or slurring) and the transformation observed under 3D rotational visualization (J-wave ⟶ slurring or slurring ⟶ J-wave). Groups *i–iii* represent the resulting phenotypic categories, and excluded cases are listed for completeness.

| 1 = Yes |
|---|
| 0 = No |

| | **Group *i*** | **V6 ECG pattern** | **J-wave ⟶ slurring** | **slurring ⟶ J-wave** |
|---|---|---|---|---|
| 1 | 017/s0053lre | J-wave | 1 | 0 |
| 2 | 040/s0219lre | J-wave | 1 | 0 |
| 3 | 045/s0147lre | Slurring | 0 | 1 |
| 4 | 048/s0171lre | J-wave | 1 | 0 |
| 5 | 049/s0173lre | J-wave | 1 | 0 |
| 6 | 052/s0190lre | J-wave | 1 | 0 |
| 7 | 057/s0198lre | Slurring | 0 | 1 |
| 8 | 083/s0268lre | Slurring | 0 | 1 |
| 9 | 099/s0387lre | J-wave | 1 | 0 |
| 10 | 110/s0003_re | Slurring | 0 | 1 |
| 11 | 165/s0322lre | J-wave | 1 | 0 |
| 12 | 173/s0305lre | J-wave | 1 | 0 |
| 13 | 180/s0374lre | J-wave | 1 | 0 |
| 14 | 198/s0402lre | J-wave | 1 | 0 |
| 15 | 240/s0468_re | J-wave | 1 | 0 |
| 16 | 241/s0469_re | J-wave | 1 | 0 |
| 17 | 245/s0474_re | J-wave | 1 | 0 |
| 18 | 248/s0481_re | J-wave | 1 | 0 |
| 19 | 249/s0484_re | J-wave | 1 | 0 |
| 20 | 250/s0485_re | J-wave | 1 | 0 |
| 21 | 252/s0487_re | J-wave | 1 | 0 |
| 22 | 258/s0494_re | J-wave | 1 | 0 |
| 23 | 259/s0495_re | J-wave | 1 | 0 |
| 24 | 273/s0511_re | Slurring | 0 | 1 |
| 25 | 277/s0527_re | J-wave | 1 | 0 |

| | **Group *ii*** | **V6 ECG pattern** | **J-wave ⟶ slurring** | **slurring ⟶ J-wave** |
|---|---|---|---|---|

| | | | | |
|---|---|---|---|---|
| 1 | 043/s0141lre | J-wave | 0 | 0 |
| 2 | 230/s0454_re | J-wave | 0 | 0 |
| 3 | 244/s0473_re | J-wave | 0 | 0 |

| | **Group *iii*** | **V6 ECG pattern** | **J-wave ⟶ slurring** | **slurring ⟶ J-wave** |
|---|---|---|---|---|
| 1 | 073/s0238lre | J-wave | 0 | 0 |
| 2 | 150/s0287lre | J-wave | 0 | 0 |
| 3 | 187/s0207_re | Slurring | 0 | 1 |

| | **J-wave/slurring but excluded** | **Reason** |
|---|---|---|
| 1 | 042/s0135lre | filiform QRS loop |
| 2 | 081/ s0264lre | filiform QRS loop |
| 3 | 109/s0349lre | Very small J-wave in V6, no J-wave in V5 |
| 4 | 186/s0293lre | filiform QRS loop |
| 5 | 203/s0424_re | filiform QRS loop |
| 6 | 227/ s0450_re | filiform QRS loop |
